# A gut pathobiont regulates circulating glycine and host metabolism in a twin study comparing vegan and omnivorous diets

**DOI:** 10.1101/2025.01.08.25320192

**Authors:** Matthew M. Carter, Xianfeng Zeng, Catherine P. Ward, Matthew Landry, Dalia Perelman, Tayler Hennings, Xiandong Meng, Allison M. Weakley, Ashley V. Cabrera, Jennifer L. Robinson, Tran Nguyen, Steven Higginbottom, Holden T. Maecker, Erica D. Sonnenburg, Michael A. Fischbach, Christopher D. Gardner, Justin L. Sonnenburg

## Abstract

Metabolic diseases including type 2 diabetes and obesity pose a significant global health burden. Plant-based diets, including vegan diets, are linked to favorable metabolic outcomes, yet the underlying mechanisms remain unclear. In a randomized trial involving 21 pairs of identical twins, we investigated the effects of vegan and omnivorous diets on the host metabolome, immune system, and gut microbiome. Vegan diets induced significant shifts in serum and stool metabolomes, cytokine profiles, and gut microbial composition. Despite lower dietary glycine intake, vegan diet subjects exhibited elevated serum glycine levels linked to reduced abundance of the gut pathobiont *Bilophila wadsworthia*. Functional studies demonstrated that *B. wadsworthia* metabolizes glycine via the glycine reductase pathway and modulates host glycine availability. Removing *B. wadsworthia* from a complex microbiota in mice elevated glycine levels and improved metabolic markers. These findings reveal a previously underappreciated mechanism by which diet regulates host metabolic status via the gut microbiota.

## Introduction

Metabolic diseases such as type 2 diabetes mellitus, hyperlipidemia, obesity, and metabolic dysfunction-associated steatotic liver disease account for over 10 million global deaths every year and result in a significant burden on the global health care system^1,2^. One attractive means of controlling these diseases is by dietary modulation^3^. Plant-based diets, such as vegan diets, have gained recent popularity for their cardiometabolic health benefits as well as for their lower environmental impact compared to omnivorous diets^4^. While vegan diets have been associated with favorable cardiometabolic health outcomes in human studies^5,6^, mechanisms underlying these benefits are complex, unclear, and require experimental investigation.

A vegan diet involves avoiding all foods derived from animals and results in consuming more plant-derived foods. Consuming a vegan diet typically results in increased consumption of dietary fiber and certain types of vitamins, minerals, and plant-derived polyphenols compared to other types of diets^7^. Increased consumption of dietary fiber has been associated with modulation of the gut microbiota, resulting in increased production of short-chain fatty acids and other beneficial metabolites^8,9^. Conversely, consuming animal products has been associated with gut microbiome mediated conversion of dietary compounds into particular metabolites, such as trimethylamine N-oxide and phenylacetylglutamine, which have been shown to potentiate cardiovascular risk^10,11^. Previous studies investigating the effects of plant-based and vegan diets on the microbiome have found associations with particular genera and species^12–15^. However, our understanding of specific mechanisms by which individual species of the microbiota might mediate putative health effects of these diets is still limited.

To address the potential role of the microbiome in mediating the healthful effects of a vegan diet, we performed analysis on a recently completed clinical trial, the Twins Nutrition Study^16^ (<u>NCT05297825</u>). One limiting factor of human trials is large inter-individual variation with regards to genetics and microbiome composition. Randomizing identical twins to either of two arms of this dietary intervention allowed us to control for some of this variation, as identical twins tend to have more similar microbiome composition in addition to having identical genetic makeup^17^. In the present study, we performed comprehensive multi-omics profiling of study participants using metagenomics, untargeted metabolomics, and immune profiling using targeted proteomics. We observed that the intervention produced generalized, diet-specific responses in each of these measurement modalities over the course of the 8-week study. Specific associations across data types allowed us to formulate hypotheses that could be tested in a reverse translational setting^18^. We used *in vitro* and *in vivo* experiments to further explore the ability for the gut pathobiont *Bilophila wadsworthia* to modulate circulating levels of the amino acid glycine and thereby influence host metabolic status. These results suggest that a vegan diet with high proportions of plant-based foods may indeed be a powerful modulator of the human microbiome and metabolome and may provide an avenue to combat non-communicable diseases.

## Results

### Participants successfully consumed nutritionally distinct diets

To investigate the effects of consuming a vegan diet on the microbiome, host metabolism and the immune system, we recruited 21 pairs of identical twins using the Stanford Twin Registry (**Table S1**). For 8 weeks, one member of each twin pair consumed a vegan diet while the other twin consumed a healthful omnivorous diet (**Figure 1A,B**). During the first 4 weeks of the study, meals were provided directly to participants to aid in diet adherence. During the second 4 weeks, participants provided their own meals with the aid of dietary counseling. Although weight loss was not discouraged, our diet design did not include a prescribed energy restriction and was not intended to be a weight loss study. Participants were told to eat until they were satiated throughout the study. At Weeks 0, 4 and 8 we collected detailed 24-hour dietary recalls, anthropometric data, blood and stool. Blood and stool were used to perform metagenomics, untargeted metabolomics and immune profiling. Preliminary results on the study design dietary patterns and clinical markers were published previously^16^. In brief, participants successfully adhered to their respective diets during both the food delivery and self-preparation phases. We observed at the end of the study that vegan diet twins had lower levels of low-density lipoprotein (LDL) cholesterol (-13.9 mg/dL, 95% CI: [-25.3 to -2.4 mg/dL]), body weight (-1.9 kg, 95% confidence interval (CI): [-3.3 to -0.6 kg]), and fasting insulin (-2.9 μIU/mL, 95% CI: [-5.3 to -0.4 μIU/mL]; **Figure S1**).

**Figure 1.**
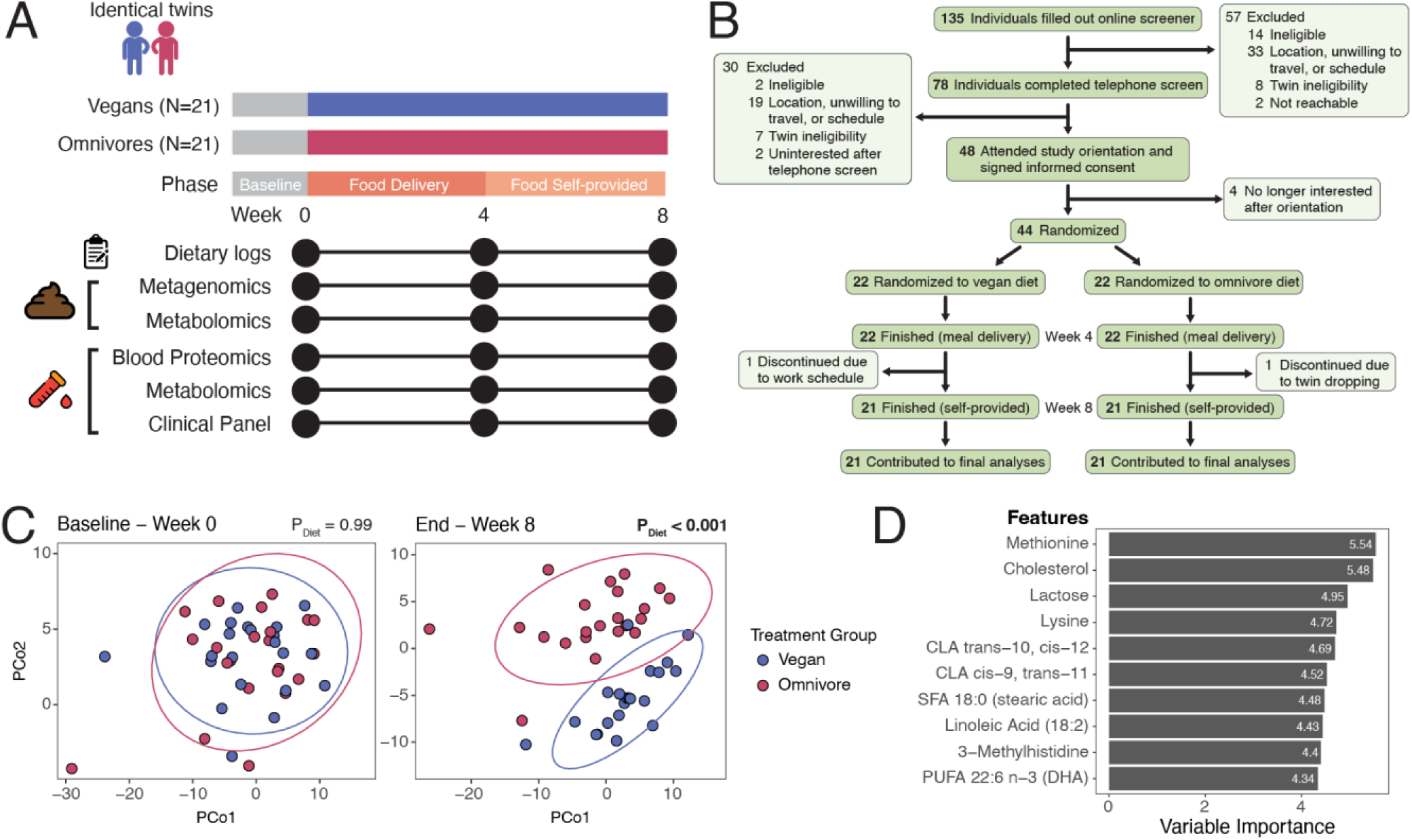
Overview of Twins Nutrition Study and implementation of vegan and omnivorous diets. (A) Trial timeline (top) and sample collection schema (bottom). Each subject was tracked for 8 weeks. Dietary recall data, blood and stool were collected at Weeks 0, 4 and 8. (B) A CONSORT Flow Diagram of the individuals recruited and screened for this study. (C) Principal coordinate analysis performed on NDSR dietary recall data at baseline (Week 0; left panel) before participants switched to their study-prescribed diet and at study end (Week 8; right panel). Dietary recall data consisted of 165 elements that described macro- and micronutrient composition of diet. Treatment group explained a significant portion of variation at study end (P < 0.001, Adonis test) but not at baseline (P = 0.99, Adonis test). (D) Ten most important variables used to predict treatment groups in a random forest model trained on NDSR dietary recall data from Week 8 diet logs. The model had a classification accuracy of 96.8% in predicting treatment groups from dietary data.

Using 24-hour dietary intake recalls processed with the Nutrition Data System for Research (NDSR)^19^ we quantified detailed components, including specific amino acids, amino acid derivatives, fatty acids, sugars, minerals and vitamins for each participant’s diets at baseline (Week 0; prior to starting the prescribed vegan or omnivorous diets) and study end (Week 8). At baseline there was no significant difference in the diets of the two groups according to micro- and macronutrient makeup (P = 0.99, adonis test, **Figure 1C, Table S2**). However, at the end of the intervention there was a statistically significant difference in the micro and macronutrient makeup of the diets (P<0.001, adonis test, **Figure 1C**). We specified a random forest model to predict a participant’s diet based solely on NDSR data. This model predicted participant diet with 96.8% percent accuracy at Week 8 compared to 39% accuracy at Week 0 (**Figure 1D**). Each of the ten most important variables identified by the model are enriched in the omnivorous diet. Among these are methionine, cholesterol, lactose and lysine, all of which are abundant components of meat and animal products that are absent or absent or lowly abundant in plant products. These results demonstrate strong adherence to their prescribed diets and highlight that adhering to vegan and omnivorous diets results in markedly different consumption of particular nutritional inputs.

### Consumption of a vegan diet resulted in a significantly different circulating and stool metabolomes

We performed untargeted metabolomics on serum and stool (1,367 and 1,531 analytes in each library, respectively) at Weeks 0, 4 and 8. We used linear mixed effect models on each analyte for each of these modalities to determine whether any of these features were enriched in the vegan or omnivorous diets. More specifically, we specified models to determine which metabolites had significant interaction effects between diet and time (that is, significantly different slopes of normalized analyte intensity between the two diets over time). For serum metabolomics, 108 metabolites were enriched in the omnivorous diet and 48 metabolites were enriched in the vegan diet (**Figure 2A**, **Table S3**). For stool metabolomics, 13 metabolites were enriched in the omnivorous diet while no metabolites are enriched in the vegan diets (**Figure 2B**). Three metabolites were significantly enriched in omnivores in both serum and stool (**Figure S2A**). Among the most enriched serum metabolites in the omnivorous diet was 3-methylhistidine (P_adjusted_ = 2.1 x 10^-5^, linear mixed effect model (LME), **Figure 2C**), a marker of meat consumption^20^. Among the most enriched serum metabolites in the vegan diet is tryptophan betaine (P_adj._ = 1.9 x 10^-7^, LME), a marker of legume consumption^21^, which the vegan group ate in large quantities. Both metabolites effectively support dietary recall data of participant adherence to their respective diets. Also among the metabolites enriched in serum for the vegan diet were microbially produced metabolites 3-indolepropionate (P_adj._ = 0.003, LME, **Figure 2C**) and lithocholate (P_adj._ = 0.016, LME)^22,23^. In the omnivorous diet serum samples, significant enrichments were observed in several uremic toxins, including 3-carboxy-4-methyl-5-propyl-2-furanpropionate (CMPF; P_adj._ = 8.7 x 10^-9^, LME) and 3-indoxyl sulfate (P_adj._ = 0.049, LME)^24,25^. We used pathway enrichment analysis^26,27^ on the metabolites enriched in both the stool and serum metabolomics data sets. At Week 8, histidine metabolism and glycerophospholipids are significantly enriched in the stool and serum, respectively, of omnivorous diet compared to the vegan diet (histidine metabolism, P_adj._ = 0.023; glycerophospholipids, P_adj._ = 4.6 x 10^-6^, global enrichment test; **Figure 2D**).

**Figure 2.**
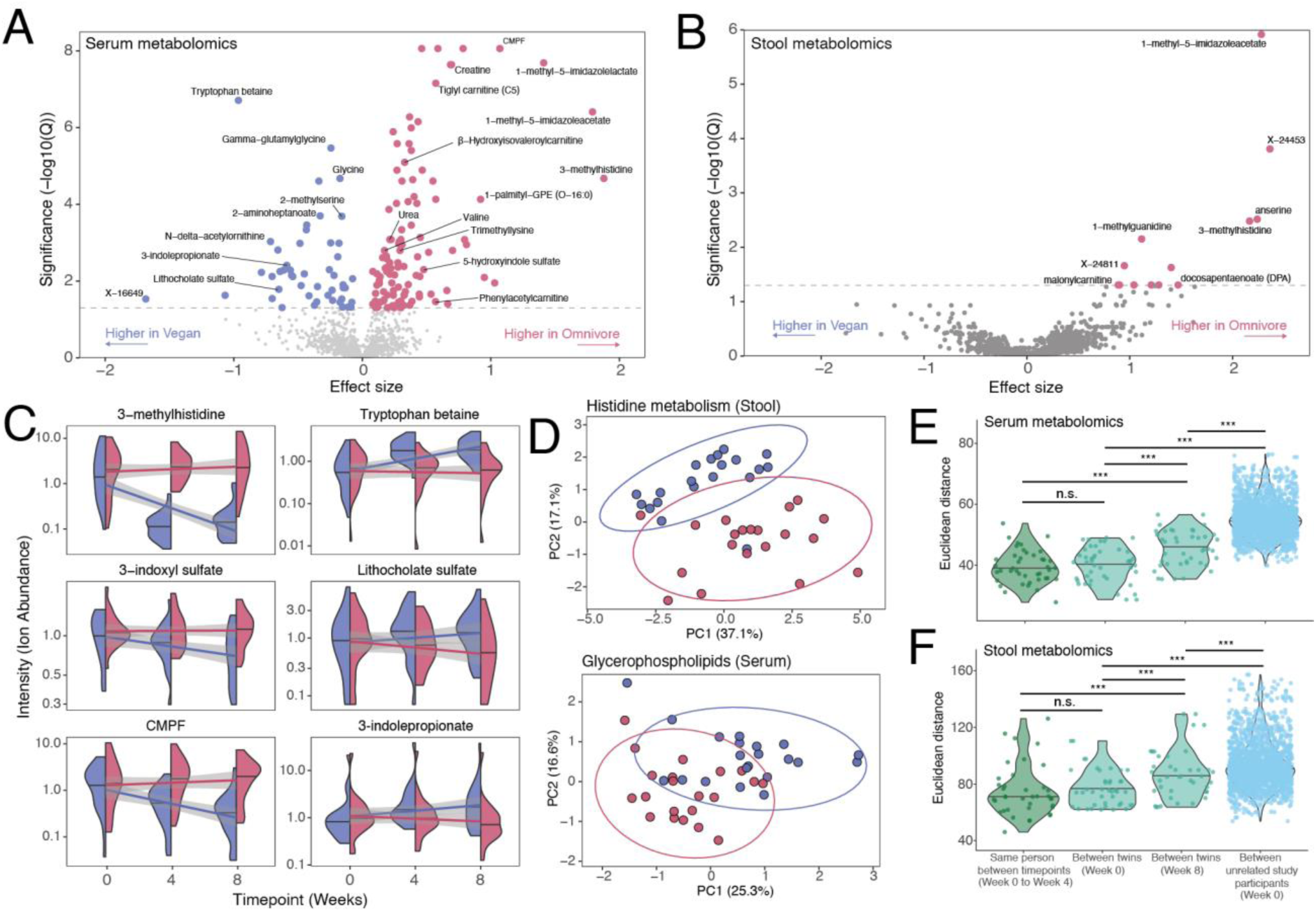
Vegan twins have distinct metabolomes compared to omnivorous twins. (A) Volcano plot of untargeted serum metabolomics showing the metabolites enriched in vegans (blue) and omnivores (red). P-values were false-discovery rate corrected with the Benjamini-Hochberg method. Horizontal dashed line represents a corrected p-value threshold of significance of 0.05. (CMPF = 3-Carboxy-4-methyl-5-propyl-2-furanpropanoic acid). (B) Volcano plot of untargeted stool metabolomics showing the metabolites enriched in vegans (blue) and omnivores (red). P-values were false-discovery rate corrected with the Benjamini-Hochberg method. Horizontal dashed line represents a corrected p-value threshold of significance of 0.05. (C) Violin plots of normalized ion intensities (arbitrary units) of a subset of serum metabolites that were significantly enriched in the vegan (blue) or omnivorous diets (red). Trend lines show linear regression fit for each diet along with the standard errors in gray. (D) Principal coordinate analysis of metabolite abundances involved in histidine metabolism in the stool metabolomics data at Week 8 (top) and of metabolite abundances of metabolites annotated as glycerophospholipids in the serum metabolomics data at Week 8 (bottom). Points are colored according to the treatment group (vegan diet = blue, omnivore diet = red). Ellipses represent 95% confidence intervals. (E) Violin plots of Euclidean distance between serum metabolomics profiles for intra-subject distance over time, between-twin distance at baseline, between-twin distance at study end and inter-subject differences at baseline. Intra-subject distances are not significantly different from between-twin distances at baseline (P = 0.21, Wilcoxon rank-sum test) but they are significantly greater than between-twin distances at study end (P = 0.00098, Wilcoxon rank-sum test). Between-twin distances at Week 0 are significantly greater than between-twin distances at Week 8 (P = 0.0038, Wilcoxon rank-sum test). Inter-subject distances are significantly greater than between-twin distances at Week 8 (P = 0.008, Wilcoxon rank-sum test). (F) Violin plots of Euclidean distance between stool metabolomics profiles for intra-subject distance over time, between-twin distance at baseline, between-twin distance at Week 8 and inter-subject differences at baseline. Intra-subject distances are not significantly different from between-twin distances at baseline (P = 0.51, Wilcoxon rank-sum test) but they are significantly greater than between-twin distances at study end (P = 2.4 x 10^-5^, Wilcoxon rank-sum test). Between-twin distances at Week 0 are significantly greater than between-twin distances at Week 8 (P = 0.00025, Wilcoxon rank-sum test). Inter-subject distances are significantly greater than between-twin distances at Week 8 (P < 2.2 x 10^-16^, Wilcoxon rank-sum test).

We leveraged the fact that our participants were identical twins to assess whether consuming vegan or omnivorous diets resulted in differences in overall metabolomic profiles between genetically identical individuals. For both serum and stool metabolomics we calculated the distance between individuals at baseline with themselves at Week 4 (“intra-subject”). We compared these values with the distance between individuals and their twins (“between-twin”) at baseline and at end-point (**Figure 2E,F**). For both data sets, intra-subject distances are not significantly different from between-twin distances at baseline (stool: P = 0.21; serum: P = 0.51; Wilcoxon rank-sum tests). However, for both serum and stool, between-twin distances at end-point are significantly greater than between-twin distances at baseline (stool: P = 0.0038; serum: P = 2.5 x 10^-4^; Wilcoxon rank-sum tests) and significantly greater than intra-subject distances (stool: P = 9.8 x 10^-4^; serum: P = 2.4 x 10^-5^; Wilcoxon rank-sum tests). This demonstrates the highly similar stool and serum metabolomic profiles of twin pairs before the intervention began, compared to unrelated subjects, and the significant difference of the profiles at the end of the dietary intervention.

We next asked which metabolites in the serum and stool metabolomics datasets had the highest and lowest between-twin variance at the end of the intervention by calculating the normalized mean absolute difference (NMAD) in metabolite intensity (**Figure S2B**). NMAD is a statistical measure used to compare the differences between paired sets of values, scaled relative to the magnitude of the values being compared. Higher metabolite NMAD values indicate higher between-twin variation and lower metabolite NMAD values indicate lower between-twin variation. For serum, 7 of the 25 most variable serum metabolites according to NMAD are plant isoflavones or phenols, such as daidzein. Additionally, 11 of these top 25 serum metabolites according to NMAD have unknown chemical identities; we speculate that these might also be plant-derived compounds. The between-twin differences in abundance of plant-derived compounds is likely reflective of markedly different intake of plant matter between the two diets. Of the 25 most similar between-twin serum metabolites according to NMAD, 11 are amino acid metabolites and 5 are lipid metabolites, compounds that, due to direct relevance to human biology, would likely be tightly regulated by the host, consistent with lower between-twin variation. For stool, 11 of the most variable 25 metabolites according to NMAD are primary or secondary bile acids. We speculate that these differences in bile acid abundance between twins might be due to broad differences in consumption of particular fatty acids between the two diets, especially with regard to higher saturated fats found in animal products^16^. Together, these findings underscore that dietary interventions can reshape metabolomic profiles even among genetically identical individuals, with plant-derived compounds and bile acids showing greater divergence between twin pairs consuming distinct diets.

### Vegan diets alter host cytokine and hormone profiles

We also performed targeted serum proteomics (70 analytes in library) using Olink on subjects’ serum in Weeks 0, 4, and 8. Two cytokines were significantly enriched in the vegan diet: stem cell factor (SCF; P_adj._ = 0.024, LME) and fractalkine (CX3CL1; P_adj._ = 0.035, LME; **Figure 3A**, **Table S4**). Both genes have pleiotropic effects on host physiology. SCF is required for maintenance of hematopoietic stem cells and erythropoiesis^28^. Recent work in mice has shown that gut microbiota-derived metabolites may influence expression of SCF in mesenchymal stem cells^29^. Fractalkine is a chemokine associated with T cell activation and monocytes. Deletion of the fractalkine gene or its receptor, CX3CR1, which is predominantly expressed in macrophages, in mouse models results in gut microbiome dysbiosis, a decrease in gut resident macrophages, colitogenic immune responses, and exacerbation of hepatic inflammation^30–33^.

**Figure 3.**
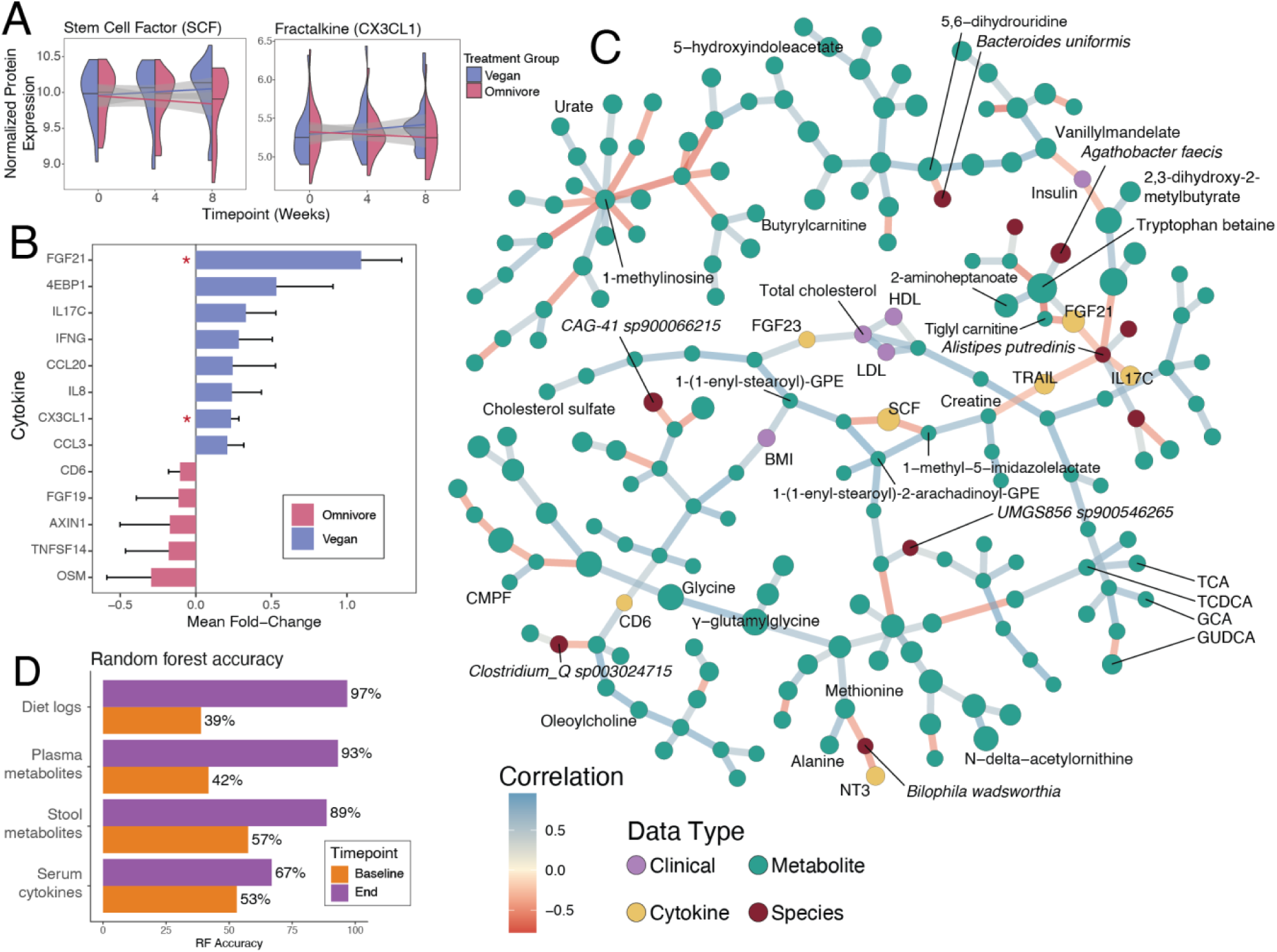
Vegan twins have distinct metabolomes and immune profiles compared to omnivorous twins. (A) Violin plots of normalized expression values (arbitrary units) of the two serum cytokines that were significantly enriched in the vegan diet: stem cell factor (SCF, P_adjusted_ = 0.024, LME) and fractalkine (CX3CL1, P_adjusted_ = 0.035, LME). Trend lines show linear regression fit for each diet along with the standard errors in gray. (B) Bar plots showing the mean fold-change between twins of the fold-change over time from baseline to the end of the study. Blue bars are elevated in the vegan twins, magenta bars are elevated in the omnivorous twins. Error bars represent the standard error. Two cytokines were significantly elevated in the vegan group (red asterisks): FGF21 (P = 0.022, Student’s t-test) and CX3CL1 (P = 0.019, Student’s t-test). (C) Correlation network graph showing significant correlations between metabolites, cytokines, clinical parameters, and gut microbial species. Nodes are colored according to data type. Edges are colored according to direction and strength of correlation. P-values of all pairwise Spearman correlations were calculated and then an adjacency matrix was assembled using only statistically significant correlations after Benjamini-Hochberg p-value correction (P_adj._ < 0.05). The backbone of this network was then extracted using the L-spar method to extract the most robust correlations. (D) Accuracy of leave-one-out cross-validation (LOOCV) of random forest models predicting treatment groups at baseline (orange bars) and study end (purple bars) for serum metabolomics, stool metabolomics and serum cytokines as well as dietary log data.

Given that genetics may play a large role in regulating the expression of this panel of proteins, we next determined which cytokines were most discordant between twins. To do this, we calculated the fold-change from Week 0 to Week 8 for each protein for each individual and then we determined the difference-of-differences for each protein according to twin pairs. This analysis revealed that fractalkine/CX3CL1 (P_adj._ = 0.019, Student’s t-test, **Figure 3B**) and fibroblast growth factor 21 (FGF21; P_adj._ = 0.022, Student’s t-test) were significantly discordant between twin pairs as a result of the dietary intervention. Both proteins were significantly enriched in the vegan twins. FGF21 is a hormone-like mediator central to metabolic homeostasis^34^. Previous work has shown that its expression increased significantly in the context of a dietary intervention that targeted the microbiota^35^. Together, these data reveal that the vegan diet exerts a significant influence on host cytokine profiles, with notable effects on SCF, CX3CL1, and FGF21. The enrichment of these proteins suggests that the dietary intervention not only impacts immune and metabolic pathways but also implicates the gut microbiome in mediating these responses.

### Vegan diets result in broad, coordinate changes to host metabolism and the immune system

In light of these broad changes to host immune and metabolic profiles, we were interested in visualizing associations between features in the different data types. We constructed a correlation network which integrated serum metabolites, cytokines, gut microbial species abundance and clinical parameters (**Figure 3C**). This analysis revealed broad, coordinated changes across these multi-omic modalities. Notably, SCF and FGF21 clustered among several gut microbial species, metabolites related to fatty acid metabolism, and clinical measures of cholesterol. Additionally, we found that the gut pathobiont *Bilophila wadsworthia* showed negative associations with a subgraph that included several proteinogenic amino acids, bile acids, and the host factor neurotrophin-3, which regulates the maintenance of peripheral and central neurons. These data suggest that shifts in diet propagate across biological domains and alter the interplay between host signaling molecules and gut microbiota constituents.

Lastly, we generated a random forest model of all participants for each of these three assays (serum proteomics, stool metabolomics and serum metabolomics; **Figure 3D**). Each model used recursive feature elimination to select the fewest number of features while maintaining the highest accuracy. As a positive control, we also used the dietary log random forest model (**Figure 1D**), which classified participants by diet arm with 96.8% accuracy (using leave one out cross-validation). Both the plasma metabolomics and stool metabolomics were nearly as accurate at predicting diet group (93% and 89% accuracy, respectively). The serum proteomics data was 67% accurate at predicting diet group. We used models generated from the baseline time point data as negative controls, which were not superior to chance at predicting diet group. These models, as well as the between-twin distance measures in **Figure 2E,F**, demonstrate that the vegan and omnivorous diets were sufficiently distinct to significantly shift the metabolomic and immunologic profiles of study participants.

### Vegans consume less glycine but have higher levels of circulating glycine

The vegan cohort consumed less dietary protein than their omnivorous counterparts (P = 6.0 x 10^-5^, Student’s t-test), mainly due to the lack of protein-dense animal products in a vegan diet. Furthermore, using dietary recall data, we observed that the vegan cohort consumed less of every amino acid than their omnivorous twins (**Figure S3A**). Curiously, the amino acid glycine was the only proteinogenic amino acid found at significantly higher abundance in serum in the vegan cohort (P_adj._ = 2.1 x 10^-5^, LME, **Figure 4A**) despite a significant decrease in dietary consumption (P = 3.05 x 10^-5^, LME; **Figure 4B,C**; **Figure S3B**). This finding of increased circulating glycine during a vegan dietary intervention study has been observed previously in humans although the mechanism behind this increase is not clear^36^. Separately, epidemiological studies have observed that increased circulating glycine is associated with protection from diabetes^37,38^, insulin resistance^39^, and other metabolic disorders^40^. Indeed, in our data, we find that increased circulating glycine is associated with lower levels of fasting insulin, which is associated with improved insulin sensitivity (P = 0.002, LME, **Figure 4D**). These data suggest that despite consuming glycine, vegans experience a counter-intuitive increase in circulating glycine which may confer metabolic benefits, including enhanced insulin sensitivity and protection against metabolic dysfunction.

**Figure 4.**
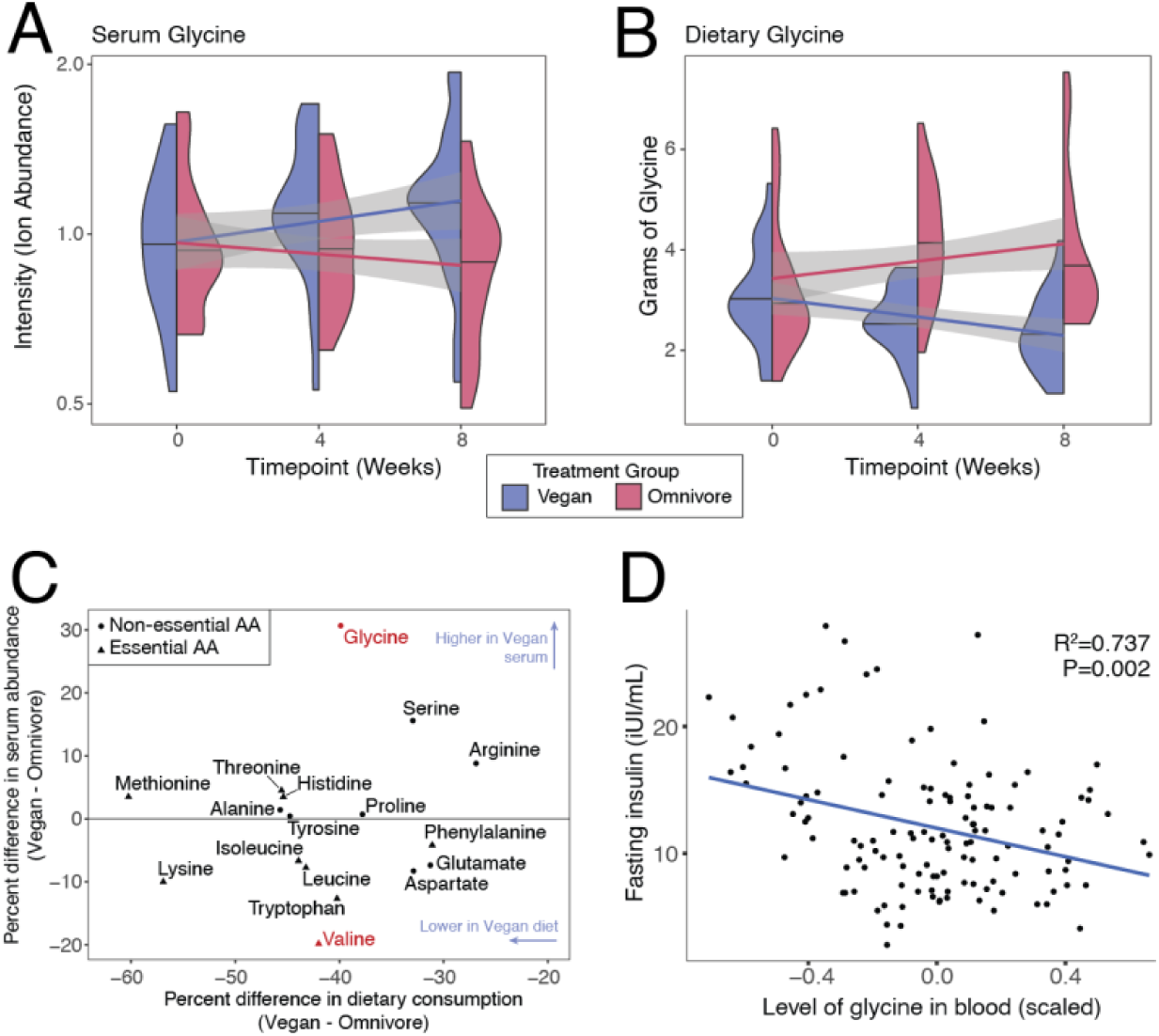
Glycine is significantly elevated in circulation of vegan twins despite lower dietary consumption. (A) Violin plot showing levels of serum glycine (normalized ion intensity) over time in the vegan (blue) and omnivore (red) treatment groups (P_adjusted_ = 2.11 x 10^-5^, LME). Trend lines show linear regression fit for each diet along with the standard errors in gray. (B) Violin plot showing levels of glycine in diet (grams) over time in the vegan (blue) and omnivore (red) treatment groups (P = 3.05 x 10^-5^, LME). Trend lines show linear regression fit for each diet along with the standard errors in gray. (C) Scatter plot comparing percent dietary consumption (x-axis) and serum abundance (y-axis) between vegan and omnivore groups of the 17 proteinogenic amino acids that were measured in both the dietary log data and serum metabolomics data. All 17 amino acids had reduced consumption in vegans; circles represent non-essential amino acids, triangles represent essential amino acids. Amino acids colored in red had statistically significant differences in serum abundance between vegans and omnivores at Week 8 (glycine: P = 0.043, valine: P =0.043, Wilcoxon rank-sum tests; multiple testing p-value correction was performed using the Benjamini-Hochberg method). (D) Scatter plot comparing normalized ion intensity of glycine (x-axis; centered and scaled) with levels of fasting insulin (international units (iU) per milliliter (mL)). Linear regression model showed a statistically significant relationship between the two variables (P = 0.002, R^2^ = 0.737).

### Vegan and omnivorous diets resulted in distinct enrichment of gut microbial species and microbial metabolic pathways

We performed metagenomic sequencing of gut microbiota from vegan and omnivorous cohorts to identify microbial taxa and metabolic pathways enriched by each diet. Linear mixed effect modeling revealed one gene significantly enriched in the vegan diet microbiome: ferredoxin-NADP+ reductase (*fpr*; *P* = 0.003, LME, **Figure 5A, Table S5**). This gene is involved in redox metabolism and regenerates NADPH to support fermentative pathways. Two genes were significantly enriched in the omnivorous diet: 3-keto-5-aminohexanoate cleavage enzyme (*kce*; *P* = 0.038, LME) and glycine reductase complex component B subunits alpha and beta (*grdE*; *P* = 0.038, LME). The kce gene is part of the lysine degradation pathway, while *grdE* participates in the glycine reductase (GlyR) pathway. Both pathways are linked to proteolytic fermentation of amino acids into short-chain fatty acids. In addition to the statistically significant genes in these pathways, both pathways show multiple additional genes enriched in the omnivore microbiomes, though just below the significance threshold (**Figure 5A**).

**Figure 5.**
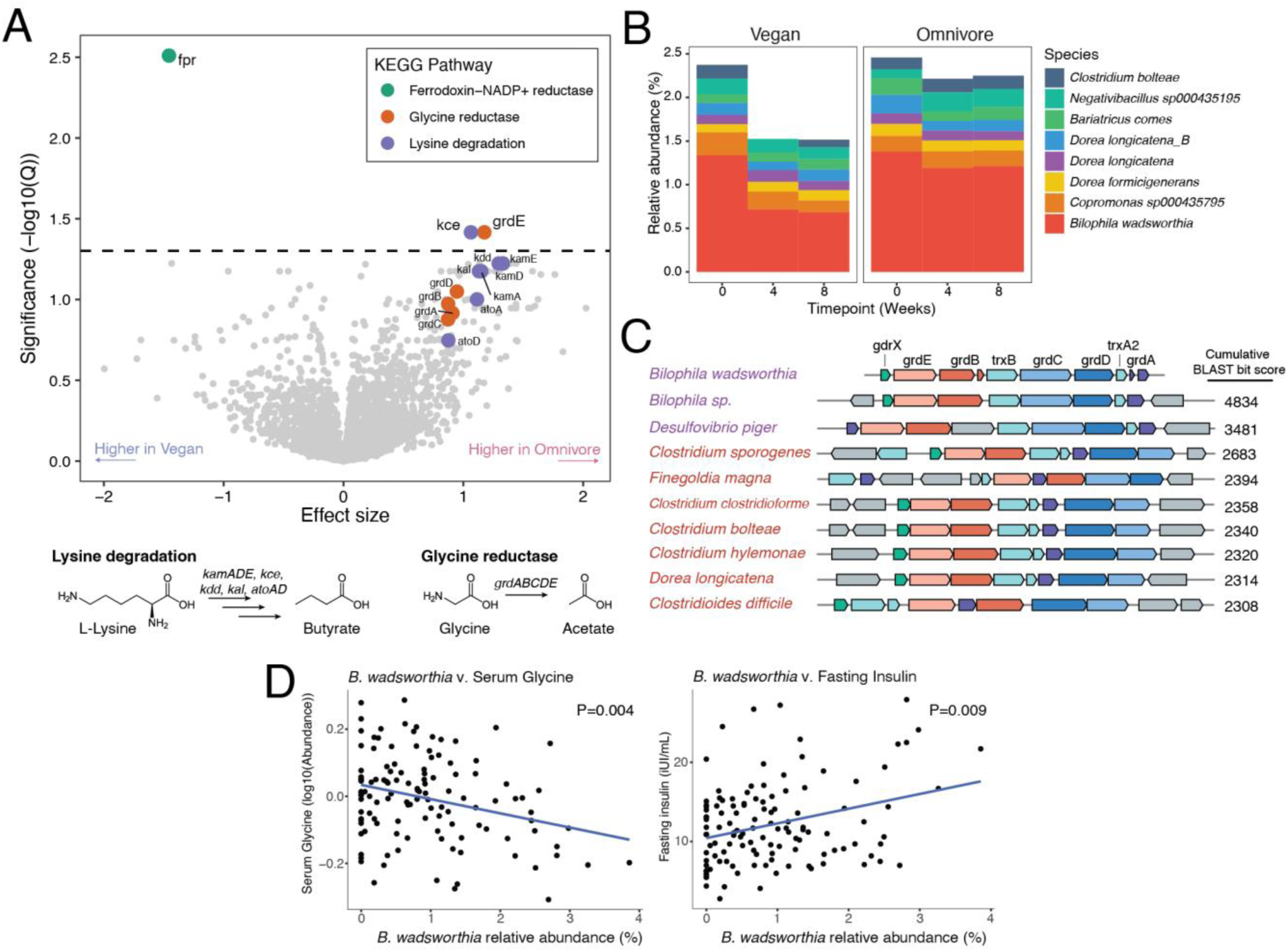
Decreased amino acid degradation in gut microbiota of vegan twins. (A) Volcano plot of KEGG orthologies significantly enriched in either vegan or omnivore diets. Genes are colored according to KEGG pathways indicated in legend. P-values were false-discovery rate corrected with the Benjamini-Hochberg method. Horizontal dashed line represents a corrected p-value threshold of significance of 0.05. (fpr = ferrodoxin-NADP+ reductase, kce = 3-keto-5-aminohexanoate cleavage enzyme, grdE = glycine reductase complex component B subunits alpha and beta). Reaction schematics below the plot demonstrate inputs and outputs of the lysine degradation and glycine reductase pathways. (B) Stacked bar chart of the relative abundance of gut microbial species that harbor the glycine reductase pathway over time and between treatment groups. (C) Multigene BLAST of glycine reductase pathway using *B. wadsworthia* as the search seed. (D) Scatter plots comparing the relative abundance of *B. wadsworthia* with normalized ion intensity of glycine(left) and relative abundance of *B. wadsworthia* with levels of fasting insulin (international units (iU) per milliliter (mL); right). Linear regression model showed statistically significant association between both comparisons (*B. wadsworthia* v. Glycine: P = 0.004; *B. wadsworthia* v. fasting insulin: P = 0.009).

Mapping these enriched genes to microbial genomes revealed their likely hosts. The *fpr* gene was most abundant in *CAG-103 sp000432375* (of the family *Oscillospiraceae*), whose abundance increased in the vegan group during the intervention (**Figure S4A**). The lysine-degrading *kce* gene was predominantly associated with *Alistipes putredinis*, which decreased in abundance in the vegan group (**Figure S4B**). The *grdE* gene of the GlyR pathway was predominantly associated with *Bilophila wadsworthia*, a member of the *Desulfovibrionaceae* family, which also decreased in abundance in the vegan group (**Figure 5B**). Examining public reference genomes, we found that the syntenic structure of the GlyR pathway of *B. wadsworthia* is strongly conserved among the proteolytic Clostridia^41^ and autotrophic *Desulfovibrio*^42^ (**Figure 5C**).

To explore the metabolic implications of these microbial genes, we correlated their abundances with serum and stool metabolites from our untargeted metabolomics dataset (**Figure S4C**). Several robust correlations emerged, highlighting potential interactions between microbial gene abundance, diet and broader metabolic patterns of the host. For example, the abundance of the oxidoreductase-encoding *fpr* gene positively correlated with redox cofactors flavin mononucleotide and nicotinic acid mononucleotide in stool. *Kce* gene abundance was positively correlated with microbially-produced, amino acid metabolites that are also uremic toxins, such as p-cresol sulfate, phenylacetylglutamine, and 3-indoxyl sulfate. *GrdE* gene abundance correlated positively with meat-derived fatty acids (e.g., docosapentaenoate) and negatively with plant-derived phytosterols, reflecting potential interactions between microbes harboring the GlyR pathway and diet. Notably, *grdE* abundance was strongly associated with secondary bile acids (e.g., 3-dehydrodeoxycholate) in stool and inversely correlated with potential inputs to the GlyR pathway in serum, namely glycine and betaine.

The identification of the microbial glycine metabolism pathway (**Figure 5A**) and its negative correlation with serum glycine was particularly striking (**Figure S4C**), given that glycine was one of the most significantly enriched metabolites in the vegan diet group (**Figure 2A**). *B. wadsworthia* is a known pathobiont linked to metabolic and immunological dysregulation^43–45^ that thrives in high fat diets^46^ and environments with higher abundance of bile acids^47^. We quantified the association between *B. wadsworthia* and serum glycine directly and found a significant negative correlation (*P* = 0.004, linear regression, **Figure 5D**). Moreover, *B. wadsworthia* abundance was positively correlated with fasting insulin levels (*P* = 0.009, linear regression, **Figure 5D**), supporting its putative role as a disruptor of metabolic health.

These findings suggest the hypothesis that the vegan diet is associated with reduced *B. wadsworthia* abundance and glycine reductase pathway activity, which in turn correlates with higher circulating glycine levels. Collectively, our results imply that *B. wadsworthia* may act as a glycine sink in the lumen of the intestinal tract, a role mitigated by adherence to a vegan diet which potentially improves metabolic outcomes.

### *B. wadsworthia* reduces glycine with the glycine reductase pathway *in vitro* via co-metabolism with pyruvate

*B. wadsworthia* has been studied extensively with regards to its unique capacity to respire taurine^48^. We wished to investigate whether *B. wadsworthia* is capable of reductive glycine metabolism given that it harbors the GlyR pathway. To date, the study of the GlyR pathway has been restricted to proteolytic Clostridia^41^ and autotrophic *Desulfovibrio*^42^. In Clostridia, glycine reduction is coupled to the oxidation of another organic substrate, a type of fermentation known as Stickland metabolism^49^. It is unknown if *B. wadsworthia* reduces glycine via a Stickland reaction and, furthermore, what the preferred oxidative Stickland partner for glycine might be.

We used a defined, minimal “freshwater salts” medium^50^ in an attempt to identify suitable growth substrates for *B. wadsworthia*. We found, as previously reported^48^, that *B. wadsworthia* grows well when taurine is the sole carbon source (**Figure 6A**). Growth is stronger when formate is added to the media as an electron donor. We also found that *B. wadsworthia* does not grow when glycine is added to the media as a sole carbon source (**Figure 6B**). Previous studies of *B. wadsworthia* demonstrated that growth was stimulated by pyruvate^51^ and early studies by Stickland demonstrated that pyruvate can be oxidized by *Clostridium sporogenes* when reducing glycine^52^. We first tested whether *B. wadsworthia* can grow using pyruvate as a sole carbon source and found that growth yield is similar to that of growth on taurine (**Figure 6C**). Growing *B. wadsworthia* in media containing equimolar glycine and pyruvate did not increase growth rate or maximal optical density (OD) compared to growth on pyruvate (**Figure 6D**). However, lack of additional growth upon the addition of glycine to medium containing pyruvate does not necessarily imply that *B. wadsworthia* is not consuming glycine or using the GlyR pathway.

**Figure 6.**
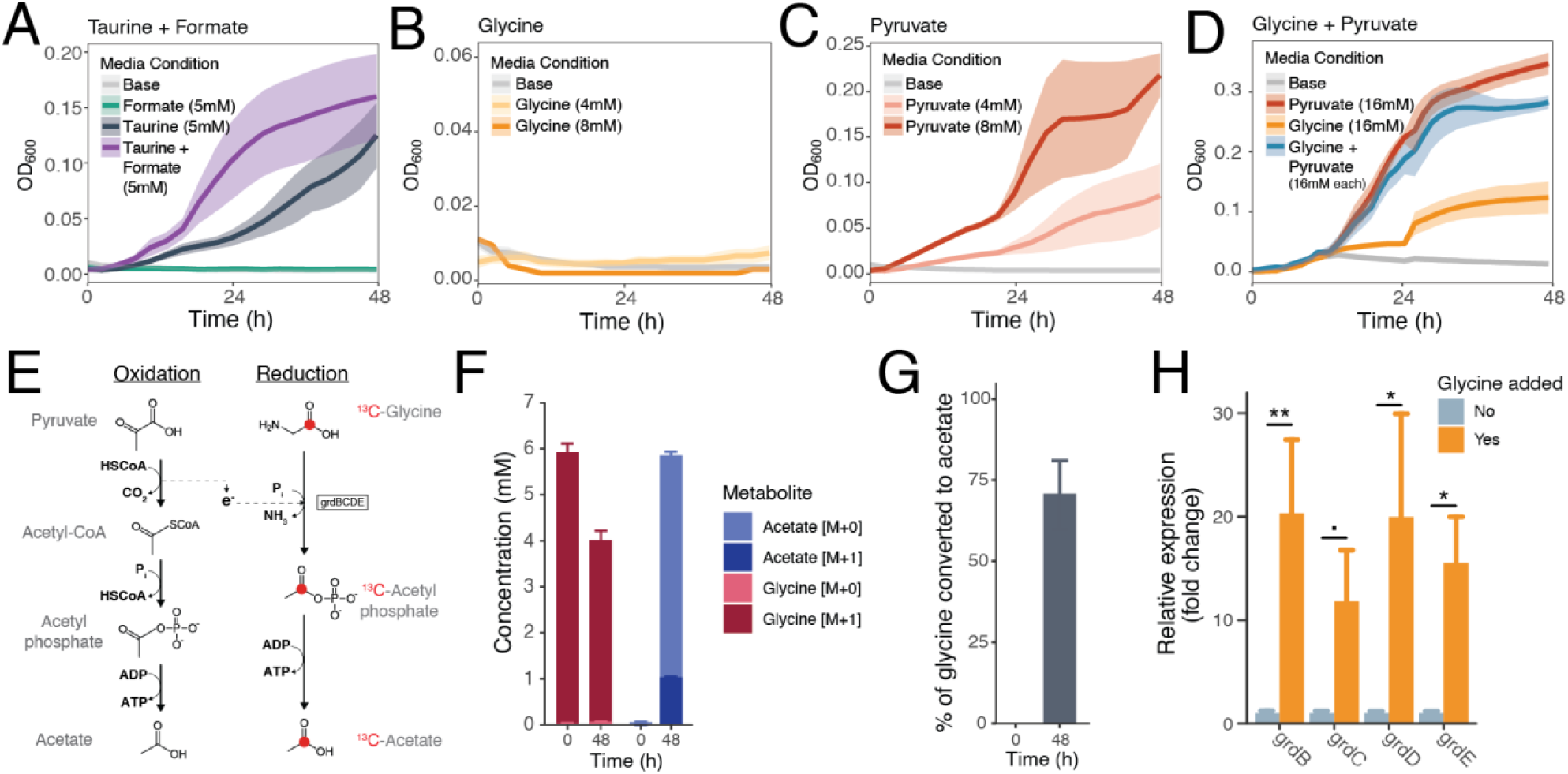
*Bilophila wadsworthia* consumes glycine via a glycine reductase pathway. (A) Growth curves of *B. wadsworthia* in liquid culture using a minimal, defined “freshwater salts” medium adding either formate, taurine, or both to the medium. Growths occurred over 48 hours. Experiments were performed in triplicate. Dots represent mean OD_600_, vertical lines represent standard error. (OD_600_ = optical density at 600nm wavelength). (B) Growth curves of *B. wadsworthia* in liquid culture using a minimal, defined “freshwater salts” medium adding varying concentrations of glycine to the medium. Growths occurred over 48 hours. Experiments were performed in triplicate. Dots represent mean OD_600_, vertical lines represent standard error. (C) Growth curves of *B. wadsworthia* in liquid culture using a minimal, defined “freshwater salts” medium adding varying concentrations of pyruvate to the medium. Growths occurred over 48 hours. Experiments were performed in triplicate. Dots represent mean OD_600_, vertical lines represent standard error. (D) Growth curves of *B. wadsworthia* in liquid culture using a minimal, defined “freshwater salts” medium adding glycine, pyruvate or both to the growth medium. Growths occurred over 48 hours. Experiments were performed in triplicate. Dots represent mean OD_600_, vertical lines represent standard error. (E) Overview of proposed Stickland reaction between pyruvate and glycine. Red dot indicates carbon-13 used to trace stable isotopes in LC-MS experiment presented in panels F and D. (F) Stacked bar charts of concentrations of glycine (M+0), ^13^C-glycine (M+1), acetate (M+0) and ^13^C-acetate (M+1) over time in defined minimal media supplemented with pyruvate and ^13^C-glycine. Experiment performed in triplicate, error bars represent standard error of the mean. (G) Bar chart showing molar ratio of ^13^C-acetate accumulation to ^13^C-glycine depletion. Error bars represent standard error of the mean. (H) Bar charts showing relative expression fold changes of genes involved in GlyR pathway with and without glycine added to the culture media. Experiments performed in triplicate, error bars represent standard error of the mean. Statistical significance was assessed using a Student’s two-tailed t test (., P < 0.1; *, P < 0.05; **, P < 0.01)

To test consumption of glycine via the GlyR pathway more directly, we grew *B. wadsworthia* in media containing ^13^C-glycine, hypothesizing that ^13^C-acetate would accumulate (**Figure 6E**). Indeed, when equimolar pyruvate and ^13^C-glycine were added to minimal media, we observed an accumulation of ^13^C-acetate over time (**Figure 6F**). When the ^13^C-acetate was expressed as a fraction of ^13^C-glycine consumed, we observed that after 48 hours nearly 75% of the carbon-13 originating from glycine was recovered in acetate (**Figure 6G**). Lastly, we performed RT-qPCR on genes in the GlyR pathway from *B. wadsworthia* cultures grown in pyruvate-containing minimal media with and without glycine. We found that each of the genes involved in the glycine reductase enzyme complex was induced (11.8-20.3-fold enrichment; P < 0.05 for 3 of 4 genes) when glycine was added to pyruvate-containing growth media compared to without (**Figure 6H**). These results demonstrate that *B. wadsworthia* can consume glycine via the GlyR pathway.

### Eliminating *B. wadsworthia* from gnotobiotic mice harboring a complex defined community results in changes to circulating glycine

We next sought to create an experimental system where we could understand whether *B. wadsworthia* alters circulating glycine *in vivo* in the context of a complex microbiota. To this end, we leveraged a recently constructed complex defined community called hCom2^53^ which uses isolates derived from human fecal samples to recapitulate the composition of a human gut microbiota. Previous work with this community has shown that single-strain dropouts can result in changes to circulating metabolite levels^54^. We performed an experiment in which we inoculated germ-free C57BL/6 with either the complete hCom2 community (“*hCom2*”; 119 strains) or the hCom2 community without *B. wadsworthia* (“*hCom2ΔBw*”; 118 strains; **Figure 7A**).

**Figure 7.**
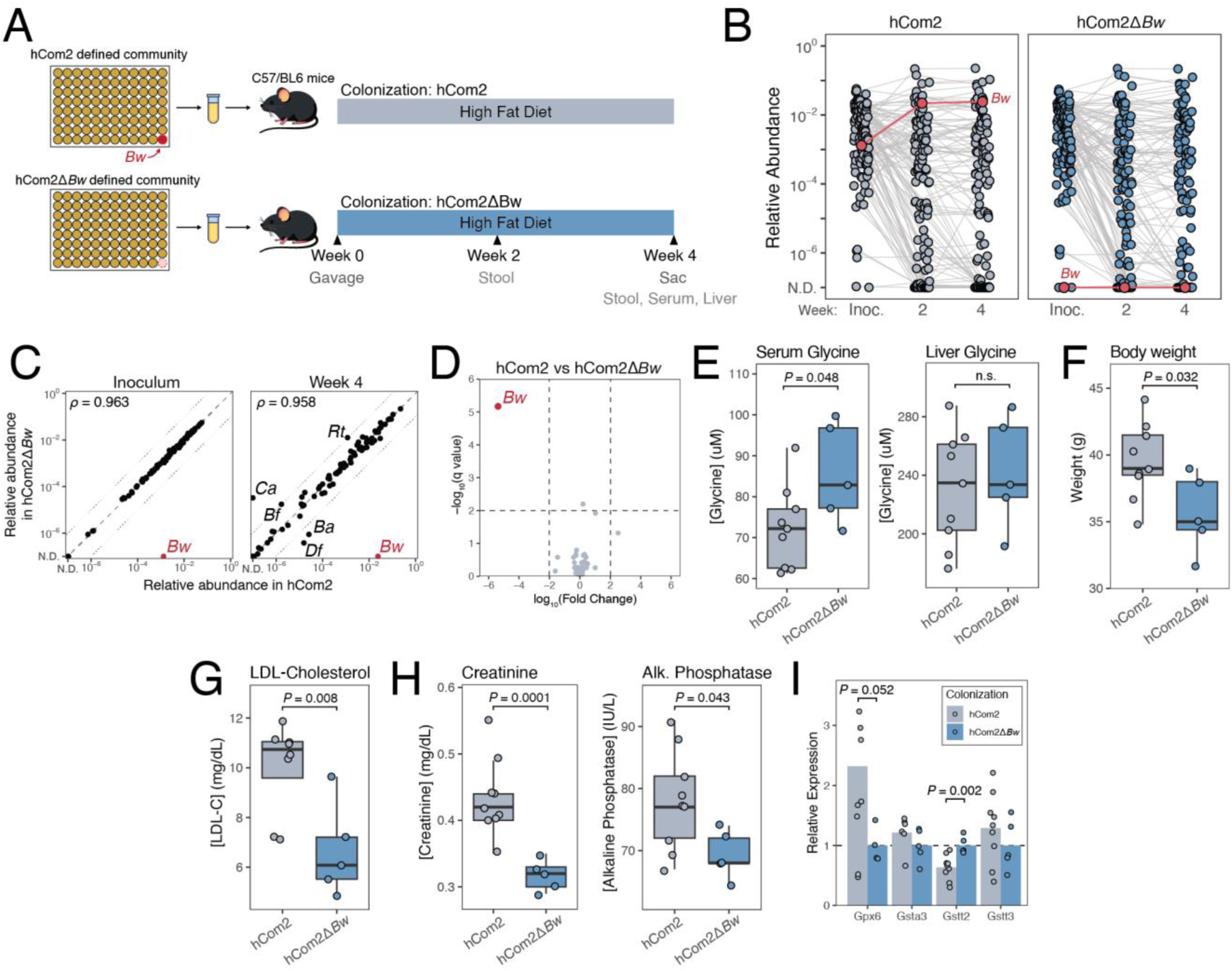
Metabolic impacts of removing *B. wadsworthia* from a complex, defined gut community. (A) Schematic of the experiment. Germ-free C57BL/6 mice were colonized with *hCom2* (n = 9 mice) or *hCom2ΔBw* (n = 5 mice) and housed for 4 weeks before sacrifice. Fecal pellets were subjected to metagenomic analysis. Serum samples were subjected to metabolite profiling and a comprehensive metabolic panel. Liver samples were subjected to metabolite profiling and gene expression profiling with RT-qPCR. (B) *B. wadsworthia* is maintained at relative abundance according to metagenomics analysis in *hCom2*-colonized mice (left) while it is undetectable in *hCom2ΔBw*-colonized mice (right). Each point is an individual species, lines connect the same species over time; the collection of dots at each time point represents the community averaged over the mice in the group. The dots and connecting lines for *B. wadsworthia* (*Bw*) are colored red. (Inoc. = Inoculum community; N.D. = Not detectable). (C) Relative abundance comparison for each species between the *hCom2* and *hCom2ΔBw* communities in the initial inoculum (left) and at Week 4 of the experiment (right). The dots for *Bw* are colored red. Pearson coefficient of correlation (*ρ*) between the two communities in the inoculum is 0.963 and at Week 4 is 0.958. Species with more than 10-fold difference are labeled (*Ca* = *Collinsella aerofaciens*, *Bf* = *Bryantella formatexigens*, *Rt* = *Ruminococcus torques*, *Df* = *Dorea formicigenerans*, *Ba* = *Bacteroides sp.*). (D) Volcano plot showing differentially enriched species between the *hCom2* and *hCom2ΔBw* communities at Week 4. Horizontal dashed line represents threshold of statistical significance. Only *Bw* was statistically significant between the groups (q-value < 0.01, fold-change > 100). Negative log_10_ fold changes indicate higher abundance in *hCom2* community. P-values were adjusted with the Benjamini-Hochberg method to account for multiple hypothesis testing. (E) Quantification of glycine in serum (left) and liver (right) using LC-MS. Serum glycine was significantly higher in *hCom2ΔBw*-colonized mice (P = 0.048, Student’s t-test). (F) Body weight was significantly higher for *hCom2*-colonized mice after 4 weeks (P = 0.032, Student’s t-test). (G) LDL-cholesterol was significantly higher for *hCom2*-colonized mice after 4 weeks (P = 0.008, Student’s t-test). (H) Creatinine and alkaline phosphatase were significantly higher in *hCom2*-colonized mice after 4 weeks (creatinine: P = 0.0001, Student’s t-test; alkaline phosphatase: P = 0.043, Student’s t-test). (I) RT-qPCR-based profiling of liver tissue for genes involved in glutathione-dependent redox homeostasis. *Gstt2* is expressed at significantly higher levels in *hCom2ΔBw*-colonized mice (P = 0.002, Student’s t-test). *Gpx6* was expressed at higher levels in *hCom2*-colonized mice, although this difference did not meet the threshold of significance (P = 0.052, Student’s t-test).

During the 4-week experiment, *B. wadsworthia* colonization was maintained in the *hCom2* mice while it was undetectable in the *hCom2ΔBw* mice (**Figure 7B**). The overall composition of these two communities was highly similar in both the initial inoculum and in Week 4 stool samples (Pearson’s correlation coefficient of 0.963 and 0.958, respectively; **Figure 7C**). In fact, *B. wadsworthia* was the only species that had a significantly different abundance between the two communities (**Figure 7D**). These results suggest that the dropout of *B. wadsworthia* from the hCom2 community does not substantially alter the overall composition of the community, which allows us to specifically assess its impact on host physiology.

We measured glycine levels in the serum of both groups of mice at the conclusion of the experiment and found that after 4 weeks, the *hCom2ΔBw* had significantly higher circulating glycine compared to *hCom2* mice (P = 0.048, Student’s t-test, **Figure 7E**). We also measured glycine concentrations in the livers of these mice and found no significant difference in liver glycine concentration between the two groups (P = 0.61, Student’s t-test). These results support the hypothesis that presence of *B. wadsworthia* is sufficient to reduce circulating glycine levels in the host.

After 4 weeks, relative to the *hCom2* mice, the *hCom2ΔBw* mice also had lower body weight (**Figure 7F**, P = 0.032, Student’s t-test) and lower low-density lipoprotein (LDL) cholesterol (**Figure 7G**, P = 0.008, Student’s t-test). We performed a comprehensive metabolic panel on serum from these mice and found that *hCom2ΔBw* mice had lower creatinine (**Figure 7H**, P = 0.0001, Student’s t-test) and lower alkaline phosphatase (P = 0.043, Student’s t-test). We performed an oral glucose tolerance test at the conclusion of the experiment but did not find statistically significant differences in the glucose response (P = 0.86, repeated measures ANOVA, **Figure S5A**) or the glucose area under the curve (P = 0.84, Student’s t-test, **Figure S5B**). It is possible that a longer duration of experiment is needed to manifest phenotypic changes as it relates to glucose metabolism.

Finally, we performed a targeted liver gene expression analysis (**Figure 7I**) to examine shifts in metabolic and detoxification pathways that might be influenced by the absence of *B. wadsworthia*. We observed changes in genes involved in antioxidant defense and xenobiotic metabolism, namely *Gstt2*, which was found at significantly higher levels in the *hCom2ΔBw* group compared to *hCom2*. These transcriptional changes suggest that removing *B. wadsworthia* from the defined community modulates not only circulating amino acid availability but also host pathways governing oxidative stress and metabolic processing.

We also performed a similar experiment using Swiss-Webster mice and a standard chow diet to test the robustness of the elevated glycine finding from the initial experiment with C57BL/6 mice (**Figure S5C**). We successfully engrafted the *hCom2* and *hCom2ΔBw* communities (**Figure S5D**). In this experiment, we also found that serum glycine was significantly higher in the *hCom2ΔBw* community relative to the *hCom2* community (P = 0.015, Student’s t-test, **Figure S5E**). We measured relative transcript expression in the liver and again found significantly higher expression of *Gstt2* (P = 0.014, Student’s t-test).

Taken together, these findings indicate that the presence of *B. wadsworthia* in the gut microbiota lowers circulating glycine levels *in vivo* and alters host metabolism and redox homeostasis. Our data highlight the importance of a specific microbial taxon within a complex community in shaping host metabolic profiles and underscore how targeted manipulation of the microbiome could beneficially modulate host physiology.

## Discussion

Understanding the mechanisms through which diet influences the gut microbiota and host metabolism is critical for designing targeted interventions to improve human health. While numerous studies have established the link between diet and gut microbiome diversity and composition, the precise mechanisms by which specific dietary components and specific microbes influence host metabolism remain elusive. Vegan diets, in particular, have been associated with favorable cardiometabolic outcomes^55–58^ and changes to the peripheral immune system^59^, yet the underlying biological mechanisms driving these benefits are not fully understood. Our study seeks to fill this gap by leveraging a randomized controlled trial of identical twins, a powerful model that allows for the control of genetic and environmental variables^60^, in which we comprehensively profile the metabolomes, microbiomes and immune profiles of study participants. By examining the impact of vegan and omnivorous diets on the gut microbiome and host metabolism, our work aims to provide a deeper understanding of the mechanism through which diet influences health outcomes.

By integrating the data from multi-omics data sets, we identified a novel link connecting the gut microbiome to host metabolism. We uncover a novel connection between vegan diets and elevated serum glycine levels, despite lower dietary glycine intake. This observation led us to investigate the role of the gut microbiota in mediating this effect, where we identified *Bilophila wadsworthia* as a key pathobiont possessing genes implicated in encoding glycine reduction. Our results are the first to demonstrate that suppression of *B. wadsworthia* in vegans is associated with higher circulating glycine levels, suggesting a potential mechanism by which vegan diets confer metabolic benefits. Furthermore, our *in vitro* studies confirm the ability of *B. wadsworthia* to conduct reductive glycine metabolism in Stickland fermentation. Highly controlled *in vivo* studies that ablate *B. wadsworthia* from a complex gut microbiome reveal that its presence not only lowers serum glycine but also affects glucose metabolism and redox homeostasis, indicating broader implications for cardiometabolic health. This study highlights the power of “reverse translation” as a means for utilizing mouse models in human-relevant mechanistic discovery of host-microbe interactions^18^.

Glycine has been repeatedly associated with positive metabolic health^61–63^. Studies in rodents, primates and humans have demonstrated that supplementing glycine in the diet helps to restore metabolic health, suggesting a causal relationship^64–67^. Additionally, previous studies have found that vegans tend to have higher levels of circulating glycine^36^. This study demonstrates a role for the gut microbiota in mediating this increase and expands upon our understanding of the multi-faceted ways in which microbial amino acid metabolism affects levels of circulating metabolites in the host which affects host metabolic and immune status^23,54,68,69^. The mechanism by which glycine might impart metabolic benefits to the host is not clear. Previous studies have identified several candidate mechanisms, including increased glutathione production (and improved redox homeostasis^64^) and glycine’s role as a neurotransmitter^70,71^. Future work will be needed to understand specific mechanisms by which glycine protects the host against metabolic pathology.

*Bilophila wadsworthia* is recognized as a gut “pathobiont” that has been shown to expand on high-fat diets and cause immunologic and neurologic pathologies in mouse models^43,45,46^. This work expands the role that *B. wadsworthia* plays in impairing host physiology in a context that is more relevant to human physiology. Additionally, recent work in mice has demonstrated that amino acid degradation by gut bacteria can drain host amino acid pools and thereby influence host metabolic health^68^. Our study expands upon this concept by highlighting a novel avenue of host-microbe amino acid crosstalk and demonstrating that this phenomenon also plays an important role in humans. This study also suggests that plant-based diets may be effective ways of reducing abundance of pathobionts like *B. wadsworthia* and thereby attenuating the harmful effects we observed in this study. Future studies will be needed to understand whether there are even more targeted ways of removing this species from the gut microbiota.

In summary, our study provides novel insights into the complex interactions between diet, the gut microbiota, and host metabolism, specifically highlighting the role of *Bilophila wadsworthia* in modulating serum glycine levels and influencing cardiometabolic health. By leveraging a randomized controlled trial of identical twins, we were able to control for genetic and environmental variables, thereby isolating the impact of vegan and omnivorous diets on the gut microbiome and host metabolome. Our findings suggest that vegan diets, by reducing the abundance of *B. wadsworthia*, may confer metabolic benefits through mechanisms involving microbial metabolism of glycine and its downstream effects on host physiology. This work not only enhances our understanding of the gut microbial pathways involved in diet-induced metabolic changes but also opens new avenues for targeted dietary interventions aimed at modulating the gut microbiota to improve human health. Future research will be essential to further elucidate the mechanistic links between specific microbial species, host metabolism, and health outcomes, potentially leading to more precise and personalized dietary recommendations that can effectively prevent and manage metabolic diseases.

## Data Availability

All data produced in the present study are available upon reasonable request to the authors. Metagenomics data is available online at https://www.ncbi.nlm.nih.gov/bioproject/1203651.

https://github.com/SonnenburgLab/Carter_et_al_analysis

## Supplemental Figures

**Figure S1.**
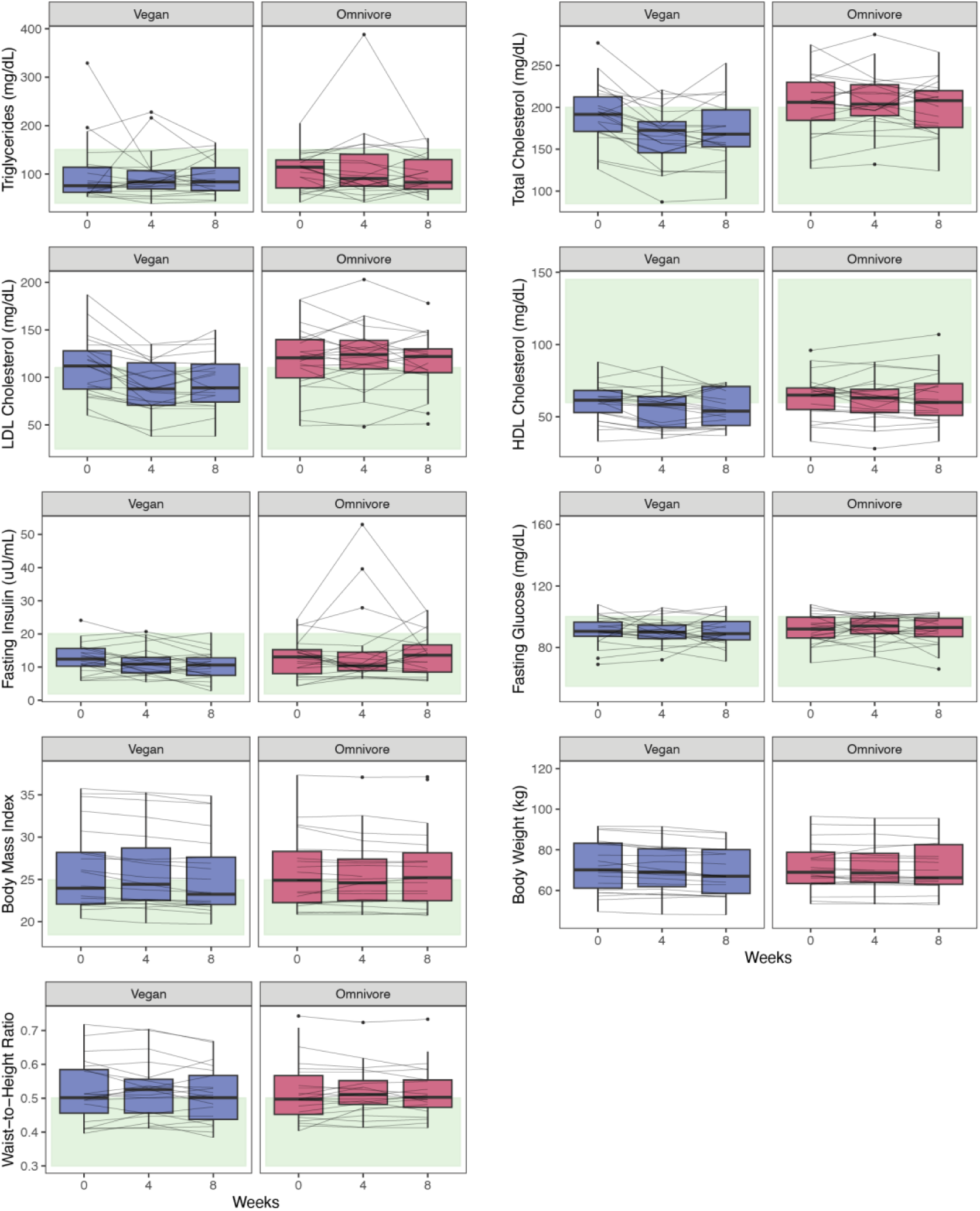
Raw values of results from standard clinical panels, related to Figure 1. Boxplots, separated by treatment group, showing triglycerides, total cholesterol, low density lipoprotein (LDL) cholesterol, high density lipoprotein (HDL) cholesterol, fasting insulin, fasting glucose, body mass index, body weight and waist-to-height ratio for each subject at 3 time points (Week 0, Week 4, and Week 8). Lines connect individual subjects over time. Green bands in the background show standard healthy reference ranges.

**Figure S2.**
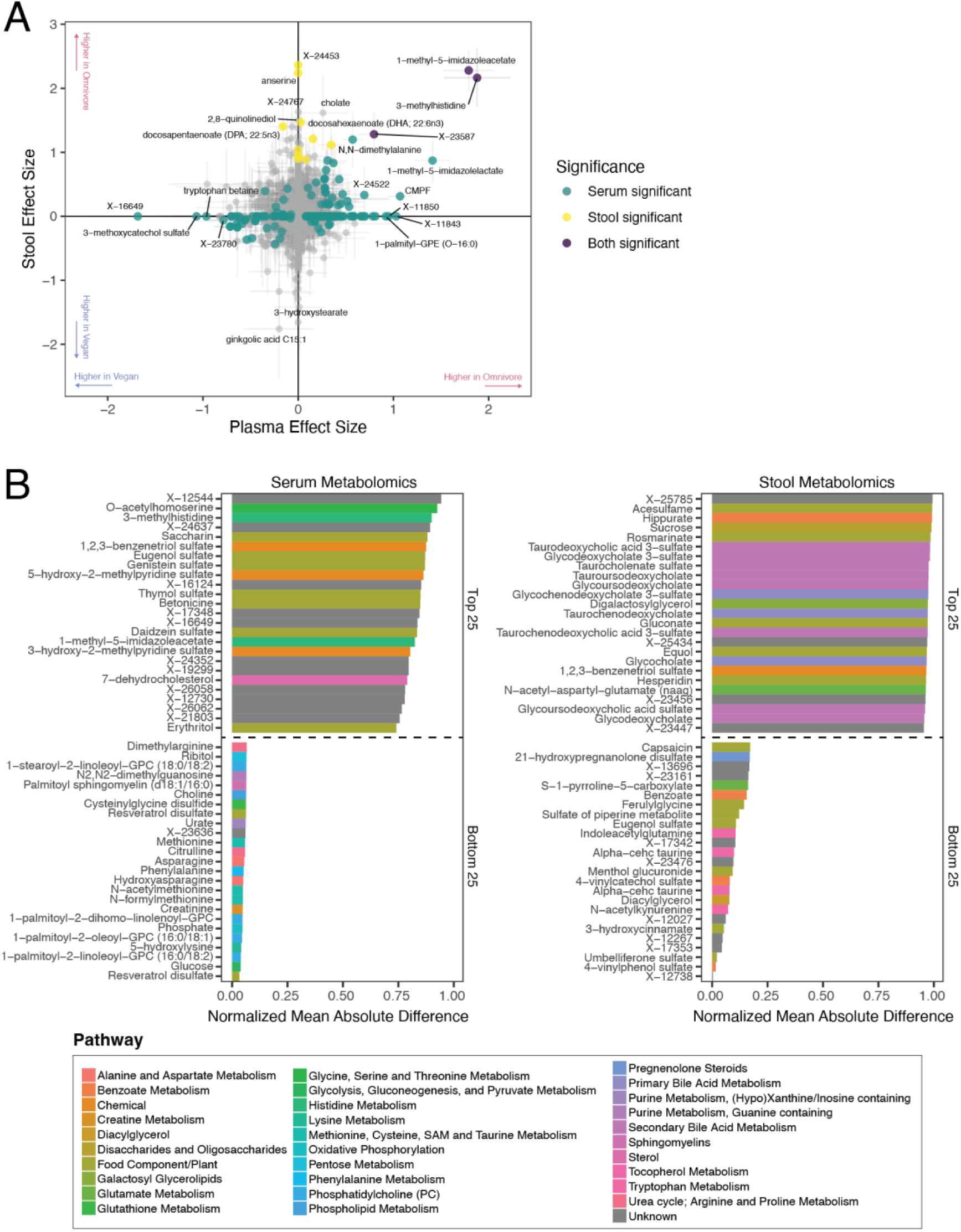
Serum and stool metabolites enriched according to treatment group or twin-based variation, related to Figure 2. (A) Effect size of enrichment of stool and serum metabolites in vegan and omnivorous diets. Same data as shown in Figure 2A and Figure 2B. (B) Top 25 and bottom 25 ranked metabolites in serum metabolomics (left) and stool metabolomics (right) according to the normalized mean absolute difference (NMAD) between twin pairs at the end of the intervention (Week 8). NMAD values reflect the extent of difference between twin pairs.

**Figure S3.**
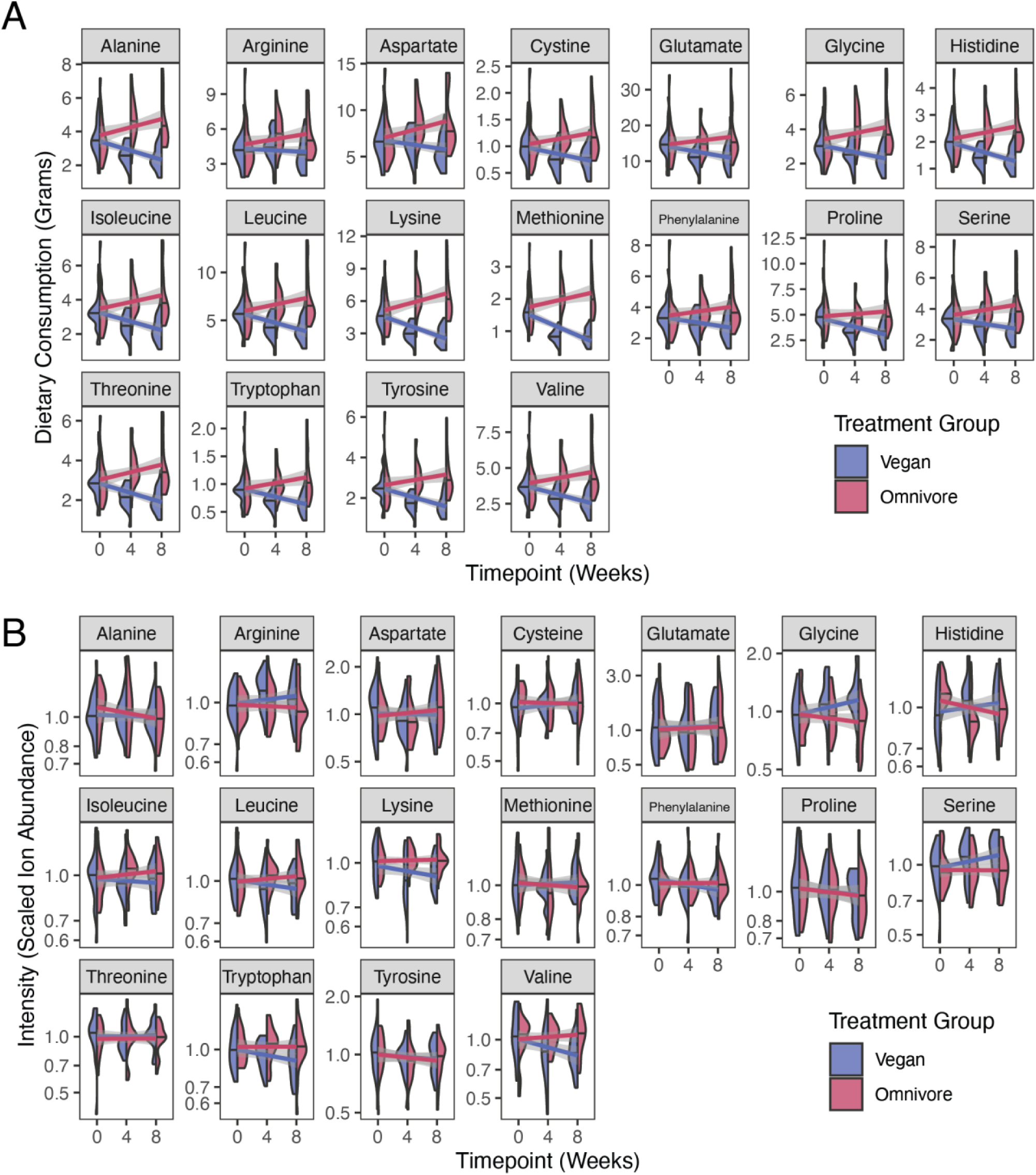
Abundances of proteinogenic amino acids in diet and serum, related to Figure 4. (A) Violin plots showing the dietary consumption of the proteinogenic amino acids over time in each treatment group. (B) Violin plots showing the serum abundance of the proteinogenic amino acids over time in each treatment group.

**Figure S4.**
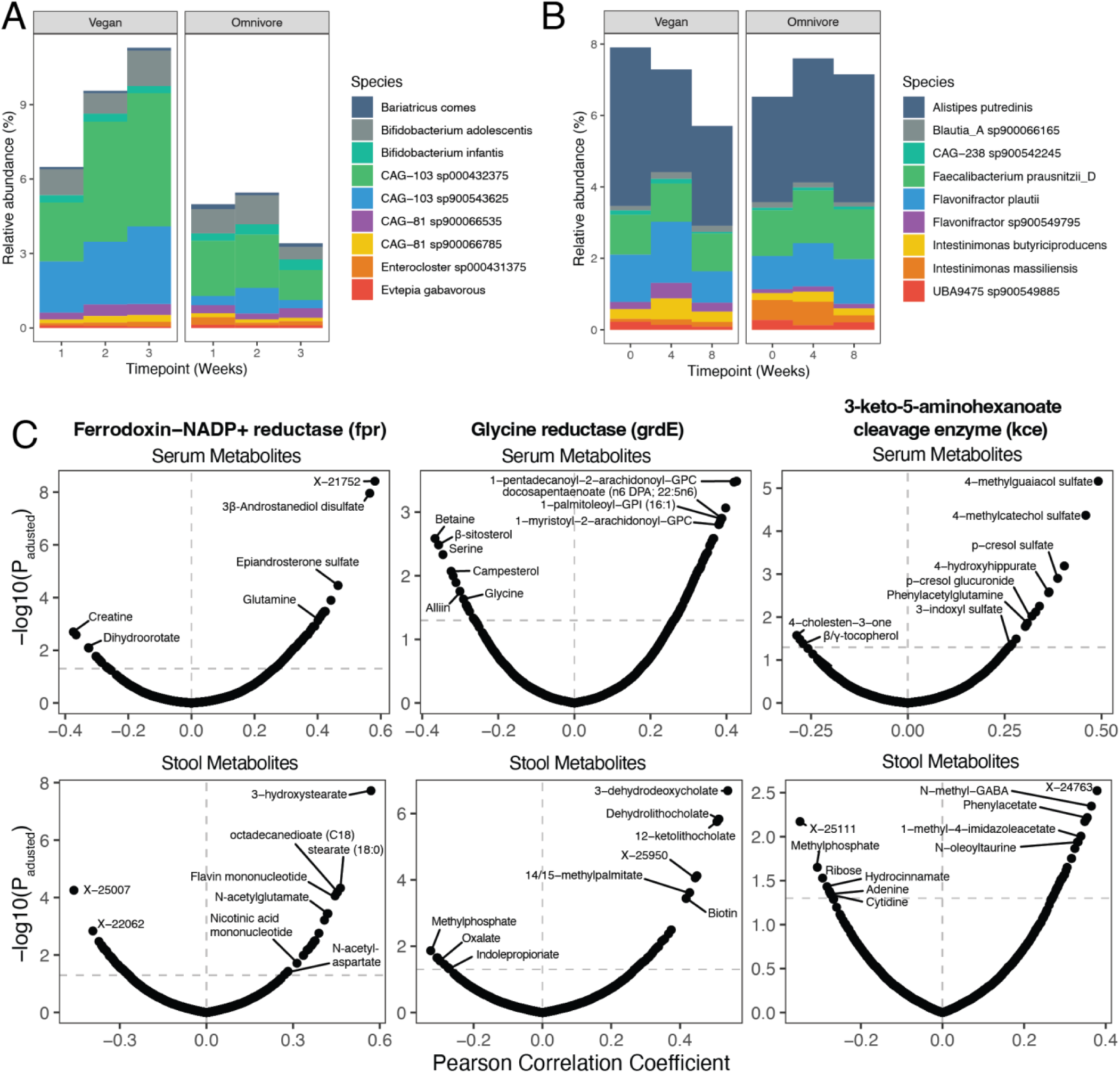
Abundance of species harboring significantly enriched genes and correlations of gene abundances with metabolite abundance in serum and stool, related to Figure 5. (A) Stacked bar plot of the relative abundances of 9 most abundant species that harbor the ferredoxin-NADP+ reductase gene (*fpr*). (B) Stacked bar plot of the relative abundances of 9 most abundant species that harbor the 3-keto-5-aminohexanoate cleavage enzyme (*kce*). (C) Volcano plots showing the Pearson correlation coefficients (x-axis) and Benjamini-Hochberg corrected p-values between the *fpr* (left panels), *kce* (center panels) and *grdE* (right panels) genes with serum (top panels) and stool (bottom panels) metabolites, respectively. Vertical dashed lines separate positive from negative correlations. Horizontal dashed lines represent respective thresholds of significance.

**Figure S5.**
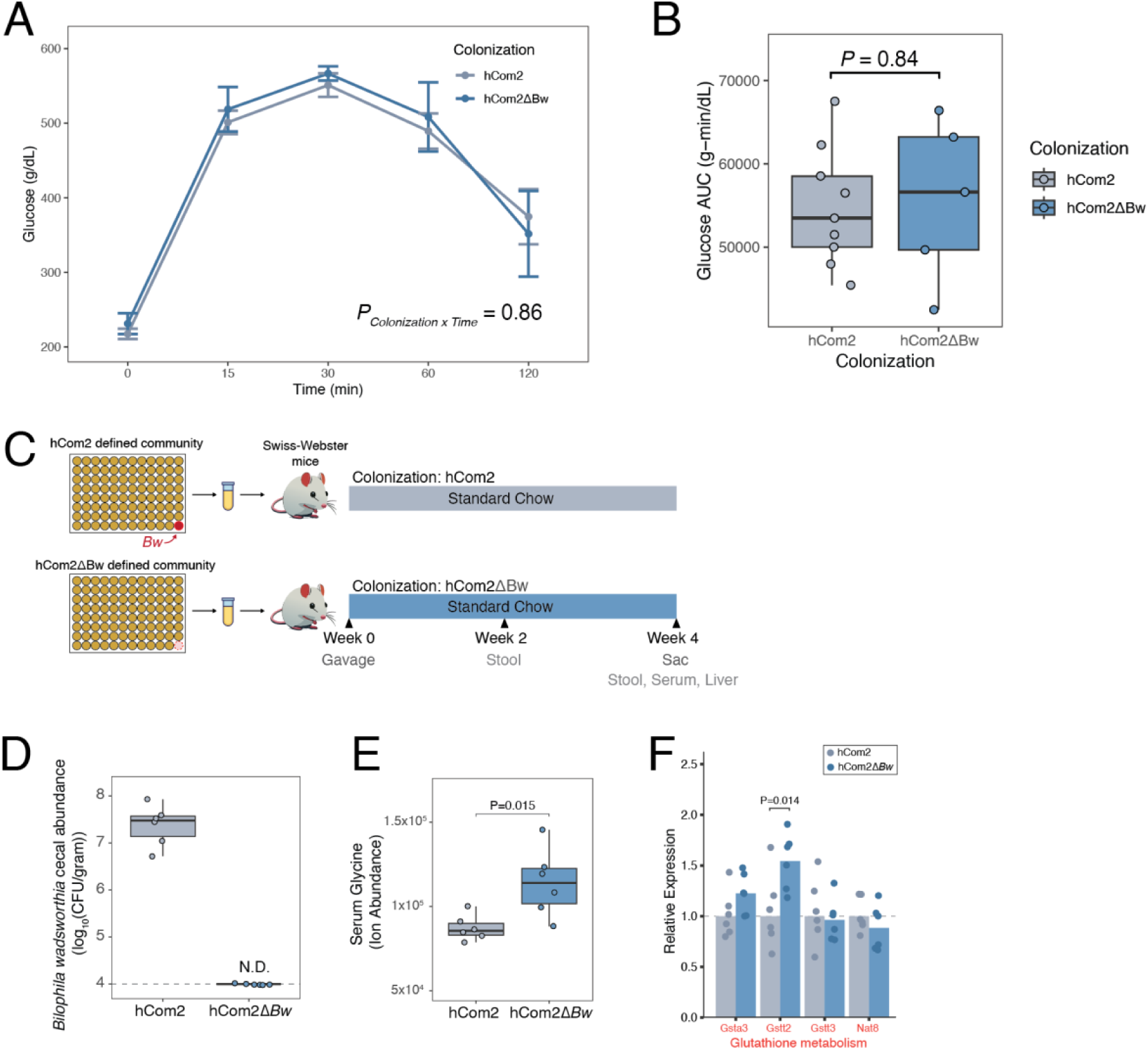
Additional data from *hCom2* mouse experiments, related to Figure 7. (A) Results of oral glucose tolerance test from experiment described in Figure 7A. Glucose clearance responses were not significantly different between groups (P = 0.86, repeated measures ANOVA). (B) Glucose area under the curve (AUC) results from data shown in (A). No difference in AUC between groups (P = 0.84, Student’s t-test). (C) Schematic of the experiment. Germ-free Swiss-Webster mice were colonized with *hCom2* (n = 5 mice) or *hCom2ΔBw* (n = 5 mice) and fed a standard chow for 4 weeks before sacrifice. Fecal pellets were subjected to RT-qPCR analysis of a marker gene for *B. wadsworthia* to quantify abundance. Serum samples were subjected to metabolite profiling. Liver samples were subjected to gene expression profiling with RT-qPCR. (D) *B. wadsworthia* is undetectable in *hCom2ΔBw*-colonized mice while being maintained at high levels in *hCom2*-colonized mice. Abundance was calculated using qPCR of the taurine-pyruvate aminotransferase gene in *B. wadsworthia*. (E) Quantification of glycine in serum using LC-MS. Serum glycine was significantly higher in *hCom2ΔBw*-colonized mice (P = 0.015, Student’s t-test). (F) RT-qPCR-based profiling of liver tissue for genes involved in glutathione-dependent redox homeostasis. *Gstt2* is expressed at significantly higher levels in *hCom2ΔBw*-colonized mice (P = 0.014, Student’s t-test).

## Supplemental Tables

**Table S1** Demographic description of study participants

**Table S2** Dietary recall data, summarized by treatment group and timepoint

**Table S3** Metadata, data and statistical results from serum and stool metabolomics

**Table S4** Metadata, data and statistical results from Olink proteomics

**Table S5** Metadata, data and statistical results from metagenomics function profiling

## Materials and Methods

### RESOURCE AVAILABILITY

#### Lead contact

All information and requests for further resources should be directed to and will be fulfilled by the Lead Contact, Justin Sonnenburg.

#### Materials availability

This study did not generate new unique reagents.

#### Data and code availability

Datasets and code for analysis are available at https://github.com/SonnenburgLab/Carter_et_al_analysis. Raw data files for metagenomics sequencing available at BioProject database, ID number: <u>PRJNA1203651</u>.

### EXPERIMENTAL MODEL AND SUBJECT DETAILS

#### Recruitment and selection of participants

Participants were recruited through the Stanford Twin Registry. The current study assessed 136 individuals for eligibility. They completed an online screening questionnaire and a clinic visit between March 28, 2022 and May 5, 2022. The primary inclusion criteria were being age 18 and older, being in general good health and being willing to consume a plant-based diet and meat, eggs and dairy. Participants were excluded if they met one of the following criteria: recent (<2 months) use of antibiotics, corticosteroids, immunosuppressive agents; a BMI ≥ 40; chronic, clinically significant, or unstable pulmonary, cardiovascular, gastrointestinal, hepatic or renal functional abnormality, as determined by medical history; Type 1 diabetes; dialysis; recent major changes in diet or lifestyle; or excessive alcohol consumption. Consort flow diagram of participant recruitment shown in **Figure 1B**. 42 participants comprised of 21 twin pairs (34 female sex and gender identifying participants (17 twin pairs), 8 male sex and gender identifying participants (4 pairs)) were used for full analysis with an average age of 39.6 with a standard deviation of 12.7 years. All study participants provided written informed consent. The objective of this study is to determine the impacts of vegan and omnivorous diets on cardiometabolic health, as well as the microbiome and immune system. The study was approved annually by the Stanford University Human Subjects Committee. Trial was registered at ClinicalTrials.gov, identifier: NCT05297825.

#### Specimen collection

Stool samples were collected at Weeks 0 (baseline), 4 (midpoint of intervention), 8 (end of intervention). All stool samples were collected in OMNIgene-GUT tubes (DNA Genotek) which enable ambient temperature storage and preservation of microbiome samples until storage at -80C when they arrived at the lab. Blood samples were collected during research clinic visits at Weeks 0 (baseline), 4 (midpoint of intervention), 8 (end of intervention). Whole blood aliquots were collected into heparinized tubes. Whole blood aliquots were incubated with Proteomic Stabilization Buffer (Smart tube, Fisher Scientific) for 12 minutes at room temperature and stored at -80C. Blood for serum was collected into an SST-tiger top tube, spun at 1,200xg for 10 minutes, aliquoted, and stored at -80C. Blood for plasma was collected into an EDTA tube, spun at 1,200xg for 10 minutes, aliquoted, and stored at -80C.

### METHOD DETAILS

#### Randomization

Participants were randomized to the vegan diet arm or the omnivore diet arm (one member of each twin pair per diet). A simple randomization was done for the groups using a random number generator (Excel), performed by a statistician not involved in the intervention or data collection.

#### Dietary recall data collection

Participants logged all their food and drink intake for 3 days (2 weekdays and 1 weekend) during Weeks 0, 4 and 8 of the study using the HealthWatch360 app. The dietitian reviewed the entries with participants to assess accuracy of entries and portions. Using the dietary log data processed with Nutrition Data System for Research (NDSR)^72^ we quantified detailed components (165 in total), including specific amino acids, amino acid derivatives, fatty acids, sugars, minerals and vitamins, for each participant’s diets at baseline (Week 0; prior to starting the prescribed vegan or omnivorous diets) and study end (Week 8).

#### Metagenomic sequencing

DNA extraction for shotgun metagenome sequencing was done using the MoBio PowerSoil kit. For library preparation, the Nextera Flex kit was used with a target of 35 ng input for 7 PCR cycles depending on input concentration. A 12 base pair dual-indexed barcode was added to each sample and libraries were quantified using an Agilent Fragment Analyzer. They were further size-selected using AMPure XP beads (Beckman) targeted at a fragment length of 450bp (350bp size insert). DNA paired- end sequencing (2×146bp) was performed on a Illumina NovaSeq 6000 using S4 flow cells (UCSD IGM Genomics Center). The average sequencing depth for each sample was 43.5 million paired-end reads (or 12.4 giga base pairs [gbp]) with a standard deviation of 20.7 million paired-end reads (or 5.9 gbp). Data quality analysis was performed by demultiplexing raw sequencing reads. BBtools suite (https://sourceforge.net/projects/bbmap/)) was used to process raw reads and mapped against the human genome (hg19) after trimming. Exact duplicate reads (subs = 0) were marked using clumpify and adapters and low-quality bases were trimmed using bbduk (trimq = 16, minlen = 55). Finally, reads were processed for sufficient quality using FastQC (https://www.bioinformatics.babraham.ac.uk/projects/fastqc/).

#### Microbiome taxonomic and functional profiling

Reads generated from each sample were mapped using Bowtie2^73^ to a recently created, comprehensive bacterial/archaeal species-level genome database^74^, and resulting mappings were processed using inStrain quick_profile (v1.2.14)^75^ and CoverM v0.4.0 (https://github.com/wwood/CoverM)). Species detected at ≥ 0.5 breadth were considered present and prevalence was calculated as the percentage of metagenomes in which the species was present. Relative abundance was calculated for each species in each sample based on the number of reads mapping to each species present divided by the total number of reads mapping to present species in a sample. Taxonomy was determined for all species-level representative genomes using GTDB (r95)^76^.

Predicted proteins were annotated from reference genomes in the reference database by first performing open reading frame prediction with Prodigal^77^ and then annotation with KEGG KO numbers using KofamScan (v1.3.0) and KOfam HMMs (v103) using default settings^78^. Annotations were only retained when the HMM score exceeded the HMM threshold. Proteins were annotated with Pfams (v32)^79^ using hmmsearch, filtered (command hmmsearch --cut_ga --domtblout), and had Pfam protein domain overlap resolved using cath-resolve-hits.ubuntu14.04 (--input-format hmmer_domtblout --hits-text-to-file).

Abundance of individual KEGG gene families per sample was calculated by first mapping reads onto the database of protein sequences described above with Bowtie2, and processing the resulting mappings and using inStrain profile (v1.2.14). Coverage was then converted to “reads per million” abundance by dividing the coverage by the total number of reads found in that sample. Then, the abundances of each protein sequence with the same KEGG orthology annotation were summed to get the abundance of each KEGG orthology in each sample.

#### Serum cytokines

Cytokine data were generated from serum samples submitted to Olink Proteomics for analysis using their provided inflammation panel assay of 92 analytes (Olink Inflammation). Data are presented as normalized protein expression values (NPX, Olink Proteomics arbitrary unit on log2 scale). We removed 22 proteins from the data set that were below the limit of detection in greater than 30% of samples.

#### Untargeted serum and stool metabolomics

Untargeted serum and stool metabolomics were performed by Metabolon, Inc. Samples were prepared using the automated MicroLab STAR system from Hamilton Company. Several recovery standards were added prior to the first step in the extraction process for QC purposes. To remove protein, dissociate small molecules bound to protein or trapped in the precipitated protein matrix, and to recover chemically diverse metabolites, proteins were precipitated with methanol under vigorous shaking for 2 min (Glen Mills GenoGrinder 2000) followed by centrifugation. The resulting extract was divided into five fractions: two for analysis by two separate reverse phase (RP)/UPLC-MS/MS methods with positive ion mode electrospray ionization (ESI), one for analysis by RP/UPLC-MS/MS with negative ion mode ESI, one for analysis by HILIC/UPLC-MS/MS with negative ion mode ESI, and one sample was reserved for backup. Samples were placed briefly on a TurboVap (Zymark) to remove the organic solvent. The sample extracts were stored overnight under nitrogen before preparation for analysis.

All methods utilized a Waters ACQUITY ultra-performance liquid chromatography (UPLC) and a Thermo Scientific Q-Exactive high resolution/accurate mass spectrometer interfaced with a heated electrospray ionization (HESI-II) source and Orbitrap mass analyzer operated at 35,000 mass resolution. The sample extract was dried then reconstituted in solvents compatible to each of the four methods. Each reconstitution solvent contained a series of standards at fixed concentrations to ensure injection and chromatographic consistency. One aliquot was analyzed using acidic positive ion conditions, chromatographically optimized for more hydrophilic compounds. In this method, the extract was gradient eluted from a C18 column (Waters UPLC BEH C18-2.1×1×00 mm, 1.7 µm) using water and methanol, containing 0.05% perfluoropentanoic acid (PFPA) and 0.1% formic acid (FA). Another aliquot was also analyzed using acidic positive ion conditions, however it was chromatographically optimized for more hydrophobic compounds. In this method, the extract was gradient eluted from the same aforementioned C18 column using methanol, acetonitrile, water, 0.05% PFPA and 0.01% FA and was operated at an overall higher organic content. Another aliquot was analyzed using basic negative ion optimized conditions using a separate dedicated C18 column. The basic extracts were gradient eluted from the column using methanol and water, however with 6.5mM Ammonium Bicarbonate at pH 8. The fourth aliquot was analyzed via negative ionization following elution from a HILIC column (Waters UPLC BEH Amide 2.1×150 mm, 1.7 µm) using a gradient consisting of water and acetonitrile with 10mM Ammonium Formate, pH 10.8. The MS analysis alternated between MS and data-dependent MSn scans using dynamic exclusion. The scan range varied slightly between methods but covered 70-1000 m/z. Raw data files are archived and extracted as described below.

Raw data was extracted, peak-identified and QC processed using Metabolon’s hardware and software. These systems are built on a web-service platform utilizing Microsoft’s .NET technologies, which run on high-performance application servers and fiber-channel storage arrays in clusters to provide active failover and load-balancing. Compounds were identified by comparison to library entries of purified standards or recurrent unknown entities. Metabolon maintains a library based on authenticated standards that contains the retention time/index (RI), mass to charge ratio (m/z), and chromatographic data (including MS/MS spectral data) on all molecules present in the library. Furthermore, biochemical identifications are based on three criteria: retention index within a narrow RI window of the proposed identification, accurate mass match to the library +/- 10 ppm, and the MS/MS forward and reverse scores between the experimental data and authentic standards.

#### Culture conditions for *B. wadsworthia*

*B. wadsworthia* isolate ATCC 49260 was used for the entirety of experiments described in this study. *B. wadsworthia* was cultured at 37 °C in a Coy anaerobic chamber using a gas mix containing 5% hydrogen, 10% carbon dioxide, and 85% nitrogen. An anaerobic gas infuser was used to maintain hydrogen levels of 3.3%. All media and plasticware were pre-reduced in the anaerobic chamber for at least 24 hours before use. The liquid media used to grow *B. wadsworthia* is the “freshwater salts” (FWS) media described previously^50^. The media contains the following components 3 g/L sodium sulfate, 0.2 g/L potassium phosphate, 0.3 g/L ammonium chloride, 0.5 g/L potassium chloride, 0.113 g/L calcium chloride, 1 g/L sodium chloride, 0.4 g/L magnesium chloride hexahydrate. After mixing with deionized water and autoclaving, the following components were added: 0.084 g/L sodium carbonate, 0.36 g/L sodium sulfide nonahydrate, 10mL/L trace mineral solution (ATCC), 10mL/L vitamins mix (ATCC), 200 ug/L 1,4-napthoquinone. Any desired media supplements were then added (i.e. taurine, formate, pyruvate, glycine, etc.). Finally, this mixture was thoroughly stirred and filter-sterilized before placing it in the anaerobic chamber to pre-reduce prior to use. All growth experiments involving quantification of OD600 were performed in an Epoch2 plate reader (Biotek) in clear 96-well Costar plates (Corning).

#### Sample preparation for LC-MS based experiments measuring glycine and acetate

To determine concentrations of glycine and acetate (as well as ^13^C-glycine and ^13^C-acetate) in culture media, mouse serum, mouse cecal contents or mouse stool, we used an LC-MS quantification method described previously^80^. Briefly, cell pellets (or some other matrix) diluted in extraction buffer containing: 80% HPLC-grade water (Fisher), 20% HPLC-grade acetonitrile (ACN; Fisher), and labeled isotopes of each SCFA/OA measured (2.5 μM d3-acetic acid [Sigma-Aldrich], 2.5 μM ^13^C-glycine [Sigma-Aldrich]). Acid-washed beads were added to the samples and the samples were subsequently shaken at 30 Hz for 10 minutes. The supernatants were then derivatized in a combination of 200 mM 3-nitrophenylhydrazine hydrochloride (Sigma-Aldrich; dissolved in 50% ACN and 50% water) and 120 mM 1-ethyl-3-(3-dimethylaminopropyl)carbodiimide hydrochloride (Pierce; dissolved in 47% ACN, 47% water, and 6% HPLC-grade pyridine [Sigma-Aldrich]). The reaction was incubated at 37°C for 30 min and subsequently diluted 50X prior to mass spectrometry analysis.

Analyses were carried out using an Agilent 6470 triple-quadrupole LC-MS and a Waters Acquity ultraperformance liquid chromatography (UPLC) ethylene-bridged hybrid (BEH) C18 column (100-mm length, 2.1-mm inner diameter, 130-Å pore size, 1.7-μm particle size). The mobile phases were water-formic acid (100:0.01, vol/vol; solvent A) and acetonitrile-formic acid (100:0.01, vol/vol; solvent B). Quantification of analytes was done by standard isotope dilution protocols. In brief, serial dilutions of the acetic acid/glycine standard solution (10 mM, 1 mM, 0.1 mM, 0.01 mM, 0.001 mM, and 0 mM) were derivatized as described above and included in each run to verify that sample concentrations were within linear ranges. For samples within linear range, analyte concentration was calculated as the product of the paired internal standard concentration and the ratio of analyte peak area to internal standard peak area.

#### RNA extraction and qPCR analysis for GlyR expression experiment

To extract RNA from *B. wadsworthia* liquid cultures, we centrifuged 5mL of culture after 24 hours of growth in FWS media plus 5mM pyruvate (with and without 5mM glycine). Pellets were re-suspended in 800uL of TRI Reagent (Zymo Research). RNA was then extracted using the Quick-RNA Fecal/Soil Microbe Microprep Kit (Zymo Research, Cat. No. R2040). RNA was quantified using a NanoDrop spectrophotometer (Thermo Fisher). cDNA libraries were generated using the iScript™ cDNA Synthesis Kit (Bio-Rad). qPCR was then performed using the qRT-PCR Brilliant III SYBR Master Mix and primers specific to the GlyR genes of *B. wadsworthia* as well as the 16S rRNA gene as a housekeeping gene control. Relative enrichment of the GlyR genes was calculated individually for each gene using the ΔΔCt method. Primers to the grd genes were designed using the NCBI Primer Blast tool.

#### Gnotobiotic mouse experiments with hCom2

##### hCom2 and hCom2ΔBw community assembly

Construction of defined, complex *hCom2* and *hCom2ΔBw* communities was performed as described previously^53,54^. Briefly, cultures of individual species were revived from frozen stocks and grown separately in their respective growth media and sub-cultured 1:10 into fresh media daily for 2-3 days. An aliquot of each culture was taken and pooled into one 1mL aliquot containing all species (in the case of the *hCom2ΔBw* community, *B. wadsworthia* was excluded from this pooling step). These mixed cultures were then pelleted in a centrifuge (5000 x g for 15 min), washed with pre-reduced sterile phosphate-buffered saline (PBS), pelleted again, and finally resuspended in 1/10 of the initial volume of a 25% glycerol/water (v/v) solution. Aliquots of the resulting synthetic communities were stored in cryovials at -80C.

##### Gnotobiotic mice

Mouse experiments were performed in either male Swiss-Webster germ-free mice (8-12 weeks of age) or male C57/BL6 germ-free mice (8-12 weeks) originally obtained from Taconic Biosciences. The experiment was performed following a protocol approved by the Stanford University Administrative Panel on Laboratory Animal Care. Mice were maintained on a high-fat chow (Teklad, TD.06414, 60% fat) and sterile water with access to food and water *ad libitum* in a facility on a 12-hour light/dark cycle with temperature controlled between 20-22 °C and humidity between 40-60%. At the start of the experiment, each mouse was gavaged with approximately 200uL of either the full *hCom2* community or the *hCom2ΔBw* community from thawed aliquots. Mice were euthanized humanely by CO2 asphyxiation and cervical dislocation after 4 weeks.

##### Sample preparation

Whole blood was collected using cardiac puncture. For each mouse, one aliquot of whole blood was used to perform a comprehensive metabolic panel at the Stanford University Animal Diagnostic Laboratory. A different aliquot of whole blood was allowed to clot at room temperature before serum separation in microtainers. Livers were harvested and flash frozen at the time of sacrifice.

##### Targeted LC-MS analysis

Liver and serum samples were subjected to targeted mass spectrometric analysis to quantify the concentration of glycine using the method described above.

##### Liver RT-qPCR analysis

We also performed RT-qPCR on RNA extracted from liver samples. RNA was extracted using Direct-zol RNA Miniprep kits (Zymo Research). RNA was quantified using a NanoDrop spectrophotometer (Thermo Fisher). cDNA libraries were generated using the iScript™ cDNA Synthesis Kit (Bio-Rad). qPCR was then performed using the qRT-PCR Brilliant III SYBR Master Mix and primers specific to glutathione^64^ synthesis genes as well as the *Gapdh* gene as a housekeeping gene control. Relative enrichment was calculated individually for each gene using the ΔΔCt method. Primers to the GlyR genes were designed using the NCBI Primer Blast tool.

##### Stool qPCR analysis

We performed qPCR on DNA extracted from stool samples to measure absolute abundance of *B. wadsworthia*. DNA was extracted using MoBio PowerSoil kits (Qiagen). qPCR was then performed using the qRT-PCR Brilliant III SYBR Master Mix and primers specific to the taurine-pyruvate aminotransferase (tpa) gene which is specific to *B. wadsworthia* (designed with the NCBI Primer BLAST tool). To generate a standard curve to quantify absolute abundance in fecal samples, we grew *B. wadsworthia* in rich media to turbidity, and plated a portion on Bacteroides bile esculin agar plates (Anaerobe Systems) to calculate colony forming units per milliliter of culture. We then extracted DNA with MoBio PowerSoil kits from this turbid media and performed qPCR on 10-fold serial dilutions.

##### Metagenomics sequencing of stool samples

Stool samples were collected during the course of the experiment. DNA was extracted from these stool samples using DNeasy PowerSoil kits (Qiagen) and the concentration of DNA was calculated using a NanoDrop spectrophotometer (Thermo Fisher). Sequencing libraries were generated in a 384-well format using a custom, low-volume protocol based on the Nextera XT process (Illumina). Briefly, the DNA concentration from each sample was normalized to 0.18 ng/μl using a Mantis liquid handler (Formulatrix). In cases where the concentration was below 0.18 ng/μl, the sample was not diluted further. Tagmentation, neutralization, and PCR steps of the Nextera XT process were performed on the Mosquito HTS liquid handler (TTP Labtech), creating a final volume of 4 μl per library. During the PCR amplification step, custom 12-bp dual unique indices were introduced to eliminate barcode switching, a phenomenon that occurs on Illumina sequencing platforms with patterned flow cells. Libraries were pooled at the desired relative molar ratios and cleaned up using Ampure XP beads (Beckman) to effect buffer removal and library size selection. The cleanup process was used to remove fragments shorter than 300 bp and longer than 1.5 kb. Final library pools were quality checked for size distribution and concentration using the Fragment Analyzer (Agilent) and qPCR (BioRad). Sequencing reads were generated using the NovaSeq S4 flow cell or the NextSeq High Output kit, both in 2×150 bp configuration.

##### Metagenomics analysis

Metagenomics analysis of *hCom2* and *hCom2ΔBw* communities was performed using a method described previously^53^. Briefly, paired-end reads were mapped to a genome database of isolate genomes that comprise the hCom2 community using Bowtie 2^73^. We then used the NinjaMap tool (https://github.com/FischbachLab/ninjaMap/releases/tag/cheng_et_al) to appropriately assign each read to a given microbial genome and then tabulate the relative abundance of each species in each sample.

### QUANTIFICATION AND STATISTICAL ANALYSIS

#### Location of Statistical Details in the Text

Results of each experiment can be found in the results and figure legends. Significant values of statistical tests are also indicated by asterisks in the figures.

#### Analysis of dietary recall data

Principal coordinate analysis was performed on the dietary log data using the vegdist function in the package “vegan” (v2.5-6), using the Euclidean distance method, and the function cmdscale from the package “stats” (v4.0.4). PERMANOVA analysis with respect to diet group at each timepoint was performed using the “adonis2” function from the “vegan” R package, with 1000 permutations.

To determine which features of the dietary logs best distinguished the vegan from omnivorous diets at the end of the intervention, a recursive feature random forest was generated using the “rfe” function from the “caret” package (using the “repeatedcv” method and 10-fold leave one out cross validation). All input parameters were first centered and scaled. These models returned the minimum number of features required for the highest accuracy (11 features to achieve a peak accuracy of 96.8%).

#### Testing for significant changes in microbe and host features during the intervention

For each of multi-omics data sets (the untargeted serum metabolomics, untargeted stool metabolomics, microbiome KEGG orthology abundance, and Olink proteomics), we constructed linear mixed effect models (LME) with Maaslin2^81^. Each model was constructed using the treatment group (“Vegan” or “Omnivore”), the time point (Week 0, 4, or 8), and an interaction term between the treatment group and the intervention phase as fixed effects. Subject identity was used as a random effect. We specified one LME per feature per data set. Features were deemed to vary significantly if the interaction term had a Benjamini-Hochberg-corrected p-value (q-value) less than 0.05. For each of the data sets we used a log2 transformation on the input data matrix as well as centering and scaling. The effect sizes shown in **Figure 2A,B** and **Figure 5A** are the coefficient terms of the interaction effect in the fit LME models.

#### Metabolomic distance calculations

In order to determine inter- and intra-participant “distances” with respect to serum and stool metabolomic profiles, we first calculated the all-versus-all distance matrices for all samples for each data set using the “vegdist” function from the “vegan” R package using the Euclidean distance method. The input data to the distance matrix calculation were the log-transformed metabolite abundances. We then subset these distances based on whether the distances were intra-subject (specifically between Week 0 and Week 4), intra-twin (at Week 0 and also at Week 8), and inter-subject (at Week 0, excluding intra-twin comparisons).

#### Metabolomic pathway ordination

In order to find host metabolism pathways enriched in either diet, the set plasma or stool metabolites enriched in either the vegan or omnivore groups (according to statistical significance of interaction effects from LMEs described above) were entered into the Enrichment Analysis software package from MetaboAnalyst^26^ (https://www.metaboanalyst.ca). We chose the KEGG pathway library to perform the global enrichment test^27^ and used the FDR-correct p-value reported by MetaboAnalyst. We then filtered the serum or stool metabolomics data sets respectively for the metabolites found in the most enriched pathways for each data set. From this filtered set of metabolites, we performed a principal component analysis on this data using the “prcomp” function from the “stats” R package (version 4.1.2).

#### Multi-omic random forest analysis

Similar to the random forest on the dietary log data described above, we also wanted to determine which features of the other multi-omics data sets (stool and serum untargeted metabolomics and Olink proteomics) that best distinguished the vegan from omnivorous diets. To do this we again generated recursive feature random forest models using the “rfe” function from the “caret” package (using the “repeatedcv” method and 10-fold leave one out cross validation). All input parameters were first centered and scaled. These models returned the minimum number of features required for the highest accuracy. In this analysis we also included the data from Week 0 (baseline) before participants started consuming their respective diets as negative controls.

#### Correlations between individual host/microbe features and clinical parameters

In order to correlate individual features from the multi-omics data sets against clinical parameters (i.e. fasting glucose, fasting insulin, etc.), we specified linear mixed effect models (using the “lmer” function from the “lmerTest” R package, version 3.1) with the clinical parameter as the dependent variable and the multi-omic feature as the dependent variable. We also included diet group (“Vegan” or “Omnivore”) as a fixed effect and participant ID as a random effect. Multi-omic features were log transformed, centered and scaled. We removed outliers where either the clinical parameter or multi-omic feature had values that were greater than 5 standard deviations away from the mean value.

#### Multi-omic correlation network

To construct a correlation network, we used the following data sets: untargeted serum metabolomics, targeted proteomics (Olink), clinical parameters and metagenomics profiles. We first calculated the log fold change from baseline to study end for each analyte for each individual. We then calculated the all-versus-all Spearman correlation coefficient (and the p-value) between all pairs of analytes using the corr.test function from the “psych” R package (v2.3.6). We then filtered the correlation matrix to rows and columns that had at least one statistically significant correlation after correcting p-values using the Benjamini-Hochberg method. We then used this adjacency matrix to construct a graph object using the “igraph” R package (v2.1.1) and then performed backbone extraction using the L-spar method from the “backbone” R package (v2.1.4) with a sparsification exponent of 0.142. The graph was then plotted and annotated using the “ggraph” R package (v2.2.1).

#### Multigene BLAST analysis

To determine which species harbored the glycine reductase pathway, we used the MultiGeneBlast tool^82^. Gene sequences of the glycine reductase operon from *B. wadsworthia* (ATCC 49260) were chosen as the search seed and NCBI’s GenBank repository as the search database. Hits were sorted according to the cumulative BLAST bit score.

### ADDITIONAL RESOURCES

Clinical trial registry NCT05297825: https://clinicaltrials.gov/study/NCT05297825

## Acknowledgements

We thank the twins for their engagement and effort to enable this study. We are deeply indebted to the Sonnenburg, Gardner and Fischbach labs for their helpful discussions as well as Christopher Dant for his useful feedback. We would like to thank Emily Ebel, Thy Nguyen and Maggie Madrigal-Moeller for technical support. We would like to thank Fredrik Bäckhed for insightful discussions about mouse experiment design. This study was funded by the Vogt Foundation and the NIH/NIDDK (R01-DK08502514, J.L.S.). C.P.W. was supported by NISH T32HL161270 from the National Heart, Lung, and Blood Institute; M.M.C. was supported by a Stanford Graduate Fellowship. J.L.S. and M.A.F. are Chan Zuckerberg Biohub Investigators.

## Author contributions

Conceptualization, C.D.G., E.D.S., and J.L.S.; patient recruitment, T.H. and J.L.R.; dietary counseling and patient interaction, D.P.; methodology and analysis, M.M.C., X.Z., C.P.W., M.L., X.M., A.M.W., A.V.C., T.N., S.H., H.T.M., visualization, M.M.C.; supervision, C.D.G. and J.L.S.; writing, M.M.C., C.D.G., E.D.S., J.L.S. All authors have reviewed the manuscript prior to submission.

